# Retinal Cytoarchitectural Alterations Across the Psychosis Spectrum and Their Correlates with Cognition: A UK Biobank Nested Case-Control Study

**DOI:** 10.1101/2025.08.18.25333916

**Authors:** Erik Velez-Perez, Cemal Demirlek, Victor Zeng, Steve Silverstein, Babatunde Aideyan, Paulo Lizano

## Abstract

Retinal structure may serve as a biomarker for psychosis-spectrum disorders (PSD) and cognition, but larger, well-controlled and detailed studies are needed. This study investigates retinal thickness differences and their association with cognition in PSD (including schizophrenia, bipolar disorder, and major depression with psychosis) compared to age-, sex-matched healthy controls (HC). In this nested case-control study using the UK Biobank data, 476 participants underwent macular optical coherence tomography (OCT). Repeated-measures ANCOVA assessed retinal thickness across two measures (left/right eyes) and two groups (PSD/HC). Comprehensive analyses were conducted, accounting for various sociodemographic (ethnicity, area-level deprivation, etc); ocular (visual acuity, intraocular pressure, etc); and health (blood pressure, body mass index) covariates, as well as excluding individuals with cardiometabolic conditions. Layers were evaluated to determine their relationship with cognition. Thinner maculae (*F*=23.02, η²p=.05, *p*<.001), ganglion cell-inner plexiform (*F*=6.42, η²p=.01, *p*=.043) and photoreceptor layers (*F*=35.31, η²p=.07, *p*<.001) were identified in PSD. The macular nerve fiber, inner nuclear, and retinal pigment epithelium layers appeared unaffected. Furthermore, smaller photoreceptor layer thickness was associated with poorer prospective memory performance (ß=0.12, B=2.15, 95% CI [0.39, 3.92], *p*=.017). The schizophrenia (*F*=26.84, η²p=.07, *p*<.001) and bipolar disorder (*F*=16.60, η²p=.05, *p*=.006) groups demonstrated the greatest as well as overlapping alterations in the photoreceptor layers. Individuals with PSD exhibit synaptic, ganglion-cell, and photoreceptor structural alterations with ocular and health-related factors —particularly cardiometabolic disorders— likely contributing to these changes. Changes in photoreceptor morphology in PSD could be related to neurobiological mechanisms associated with visual processing and memory deficits.

## 1. Introduction

Psychosis spectrum disorders (PSD) – including schizophrenia (SZ), bipolar disorder (BD), and major depressive disorder with psychosis (MDD-P) – are characterized by psychosis, mood disturbances, cognitive impairments, functional decline, and medical comorbidities [1,2]. PSDs share phenotypic and neurobiological features, justifying a transdiagnostic approach to understanding shared mechanisms [3,4].

An emerging area in PSD research involves the visual system, which may offer novel insights into visual cortical [5–7] and visual processing [8,9] deficits. As the first relay in the visual pathway, the retina consists of a highly organized network of neurons that transmit signals to the visual cortex. The retina has developmental and structural similarities to the brain, rendering it biologically informative and accessible for investigation [10,11]. Optical coherence tomography (OCT), a non-invasive imaging modality, enables precise quantification of individual retinal layers (***Figure 1A-C***) [12,13] and facilitates the detection of cytoarchitectural changes. These retinal changes may reflect broader neurobiological alterations associated with PSD, with the advantages of low cost, scalability, and minimal participant burden—positioning it as a promising tool for large-scale research and potential clinical translation [10–12,14–17].

**Figure 1.**
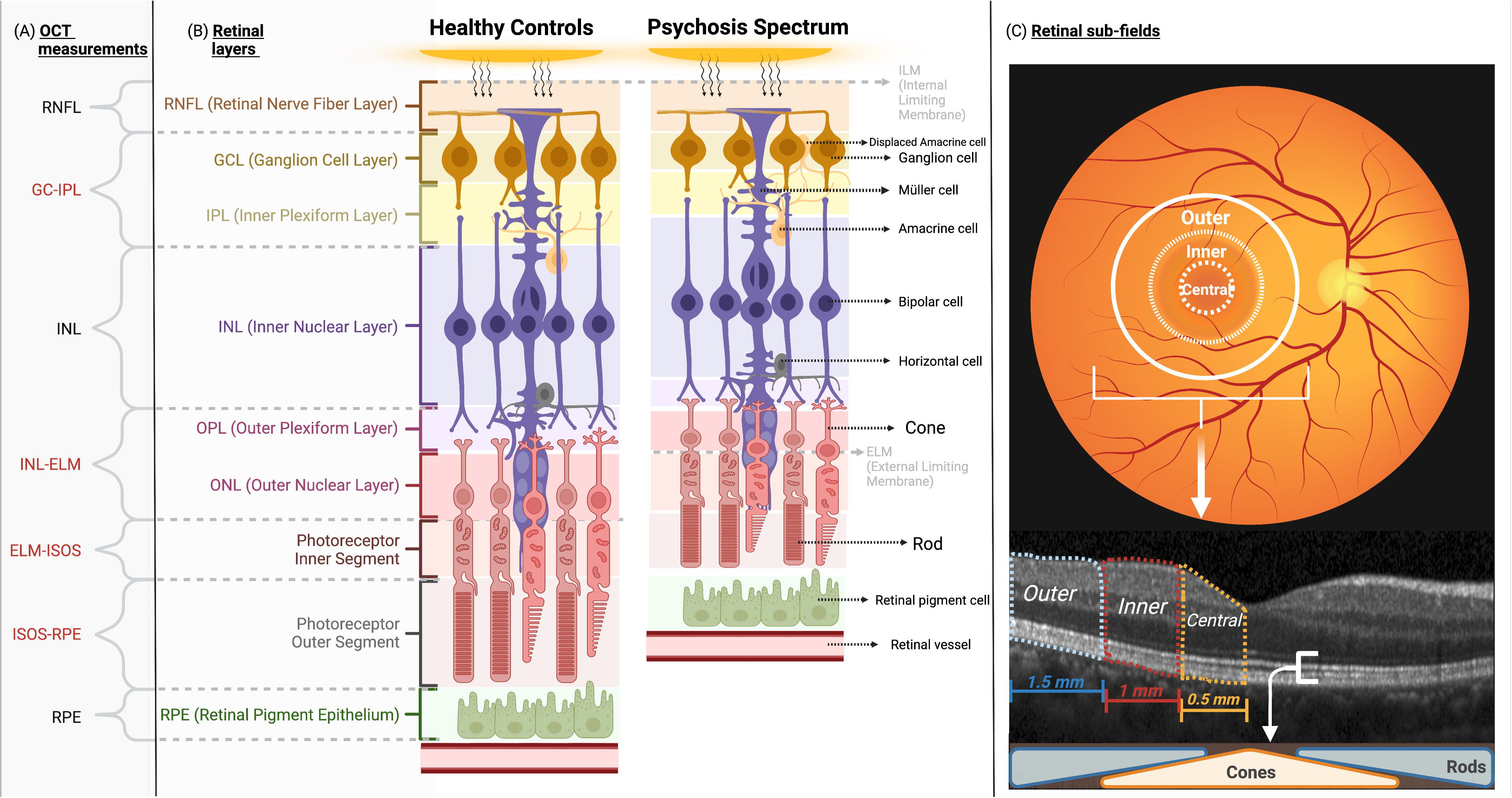
This illustration depicts three key aspects: (A) layers as determined by spectral domain optical coherence tomography (OCT) measurements and its correspondence with the specific retinal layers assessed in the UK Biobank dataset. Affected layers in PSD—GC-IPL, INL-ELM, ELM-ISOS, and ISOS-RPE—are labeled in red, (B) the arrangement of retinal layers and their associated cells, along with a representative (not an exact spatial alignment) image showing lower thickness of certain layers in PSD, and (C) the delineation of retinal sub-fields. Of note, rods populate the outer sub-field, whereas cones predominantly reside in the central sub-fields. In both panels A and B, the lower portion of the figure denotes the posterior (outer) aspect of the retina. The macula layer encompasses all layers from the RNFL to the RPE. RNFL (Retinal Nerve Fiber Layer); GC-IPL (Ganglion cell and Inner Plexiform Layers); INL (Inner Nuclear Layer); INL-ELM (Inner Nuclear Layer to External Limiting Membrane); ELM-ISOS (External Limiting Membrane to photoreceptor Inner Segment / Outer Segment); ISOS-RPE (photoreceptor Inner Segment / Outer Segment to Retinal Pigment Epithelium); RPE (Retinal Pigment Epithelium layer).

Recent meta-analyses of OCT studies have identified structural retinal alterations in PSD. A meta-analysis of 1,956 SZ, 1,189 BD, and 3,135 healthy eyes reported thinner macular ganglion cell–inner plexiform (GC-IPL), outer nuclear (ONL), and retinal pigment epithelium (RPE) layers in PSD. However, no differences were found in the retinal nerve fiber layer (RNFL) [18]. In contrast, another meta-analysis found RNFL thinning in SZ and BD but not in MDD [19]. While these findings support retinal alterations in PSD, most studies have focused on inner layers, leaving outer layers—including photoreceptors—largely unexplored. This is a notable gap given the high metabolic demands and vulnerability of photoreceptors to oxidative stress and mitochondrial dysfunction, both of which have been implicated in psychosis [20,21].

Moreover, prior studies have relied on small samples and lack consistent adjustment for important confounding variables. Meta-analytic evidence suggests that retinal thickness is influenced by age, sex, illness duration, medical comorbidities, and study quality [18,19,22]. Other factors, such as cardiometabolic disorders, visual acuity, intraocular pressure, and social determinants of health—particularly ethnicity and area-level deprivation—have also been linked to retinal thinning [23–32]. Non-White individuals and those with greater social deprivation scores consistently show smaller macular thickness, emphasizing the importance of comprehensive covariate control in population-based studies [24,25,31].

Genetic studies using the UK Biobank (UKB) have identified associations between retinal thinning and polygenic risk for SZ, as well as single-nucleotide polymorphism associations with retinal structure [33,34]. Additionally, electroretinography studies have shown cone and rod waveform anomalies in unaffected youth at familial risk for psychosis spectrum [35–37]. These findings support a genomic basis for retinal alterations in psychosis and underscore the utility of large-scale datasets like the UKB for disentangling these effects from confounding influences.

In addition to structural alterations, retinal markers may reflect functional brain outcomes. Cognitive deficits are core features of PSD, often emerging before the onset of psychosis and contributing to long-term disability [38,39]. Prior studies with small and medium-sized samples have linked retinal changes to lower cognitive performance [6,11,14,40–42], suggesting that the retina may be a surrogate marker for cognitive dysfunction.

This study represents an effort to address prior limitations by investigating retinal cytoarchitectural differences across the full psychosis spectrum using a transdiagnostic, nested case-control design within the UKB. We compare individuals with PSD to matched healthy controls (HCs), adjusting for sociodemographic, clinical, and ocular confounders. Our primary objective is to characterize layer-specific and subfield-specific retinal differences. Our secondary aim is to examine associations between retinal morphology and cognitive functioning. We hypothesize that PSD will be associated with retinal alterations and that these changes will correlate with impaired cognition.

## 2. Methods

### Design, Participants, and Setting

A nested case-control study was conducted using the UKB, a prospective cohort of ∼500,000 individuals aged 40-69, recruited across the United Kingdom between 2006-2010 [43]. The study followed the Declaration of Helsinki and was approved by the Northwest Multicenter Research Ethics Committee (approval number: 11/NW/0382). All participants provided written informed consent; individuals who later withdrew consent were excluded. Reporting followed the STROBE and the RECORD guidelines [44,45].

PSD cases were identified using the ICD-10 codes, including SZ (F20–F29), BD (F30.2, F31), and MDD-P (F32.3, F33.3). Co-occurring psychiatric diagnoses were permitted, but self-reported psychiatric diagnoses were excluded to minimize misclassification. HC participants were required to have no documented mental or behavioral disorders (ICD-10 Chapter V).

Exclusion criteria for both groups included diagnoses of eye and adnexal disorders (Chapter VII), diseases of the nervous system (Chapter VI), and other severe systemic illnesses.

Participants with duplicate entries, retinal imaging in only one eye, missing age or birth sex, or retinal thickness outliers (Z-score >3 or <-3) were excluded. Data “instances” were matched when available. To minimize confounding for age and sex, and to reduce the risk of over interpretation that can occur when using larger non-psychiatric samples for comparison, PSD and HC participants were frequency-matched 1:1 on age and sex using the case-control matching function “Match Cases to Controls” in IBM SPSS v.29.0.2—i.e., each PSD participant was paired with an HC of the same sex and age.

### Variables

The dependent variables included macula-centered retinal scan measures using the Topcon 3D OCT 1000 Mk2 device (Topcon, Inc., Oakland, NJ) for both eyes under mesopic conditions and without pupil dilation. This scanning technique involved 512 A-scans per B-scan with 128 horizontal B-scans arranged in a 6 × 6-mm raster pattern. The macular scans were segmented using Version 1.6.1.1 of the Topcon Advanced Boundary Segmentation into total macular retina (mRetina) and eight layers, including mRNFL, mGC-IPL, inner nuclear layer (mINL), INL to retinal pigment epithelium (mINL-RPE), INL to external limiting membrane (mINL-ELM), ELM to photoreceptor inner/outer segments (mELM-ISOS), ISOS to RPE (mISOS-RPE), and RPE (mRPE) (***Figure 1A-C***) [46]. As recommended, Data Fields 28552/28553 with image quality score >45 were used for OCT image quality control [47].

Secondary exposure variables and covariates included sociodemographic factors (age, sex, ethnicity, annual household income, and Townsend Deprivation Index [TDI]) [48], health status indicators (body mass index [BMI], systolic blood pressure), behavioral factors (tobacco smoking exposure, alcohol consumption), ocular health indicators (best-corrected visual acuity [BCVA] in logMAR, corneal hysteresis, intraocular pressures, spherical equivalent refractive error), and cognitive performance (processing speed via Reaction Time [Snap] test, fluid intelligence via Intelligence Test, and prospective memory via Prospective Memory [Shape] test) [49,50]. Several other cognitive tasks in UKB were not included since they had substantial missingness (>50%) and was administered to smaller sub-samples.

### Statistical analysis

Group differences in OCT and sociodemographic, health, and ocular indicators were first assessed between PSD and HC using χ² or Fisher-exact tests and independent-sample t-tests. To identify covariates influencing retinal thickness, linear regression models adjusted for age and sex were used to evaluate associations between these variables and retinal layer thickness, as done in other UKB studies [25,42,51]. Mediation analyses utilizing linear regression and the Sobel test examined whether TDI mediated the associations between ethnicity or income and PSD status to determine the effects of ethnicity and income on retinal thinning. Variables associated with lower retinal thickness in ≥2 layers were included in Model 2.

Group differences in retinal layer thickness were examined using repeated-measures ANCOVA, using 2 measures (left/right eyes) X 2 groups (PSD/HC). Before ANCOVAs, model residuals were inspected (skewness=0.11/0.12 and kurtosis=0.22/0.25 for all retina/cognition, respectively), indicating normality. Homogeneity of variances was confirmed with Levene’s tests for retinal layers (all p>0.05). Model 1 adjusted for age and sex. Model 2 additionally adjusted for TDI, BMI, BCVA, corneal hysteresis, and spherical equivalent refractive error. Model 3 excluded PSD participants with cardiometabolic disorders, since the exclusion steps and 1:1 matching, incidentally yielded a HC sample without CMD. Post hoc ANCOVA analyses were conducted for significant layers. To control for multiple comparisons, raw p-values were adjusted with SimpleM (*q*)—which accounts for correlation among layers [52]. First, subfield-level analyses (central, inner, outer) were performed to examine regional thinning patterns; subfield data were unavailable for RNFL and GC-IPL [11]. Second, diagnostic subgroup (SZ, BD, BD-P, MDD-P) differences were evaluated based on prior evidence suggesting greater thinning in SZ [18,19].

In secondary analyses, linear regressions adjusted for age and sex were used to examine associations between cognition (reaction time, fluid intelligence, prospective memory) and all retinal layers. Mediation analyses were performed for significant associations to determine whether PSD status mediated the relationship between cognition and retinal morphology for exploratory purposes, assuming the directional ordering cognition → PSD → retina on theoretical grounds, given evidence that cognitive impairments often precede the onset of psychosis [1,2,31]. Additionally, indirect effects were re-evaluated with 5000 bias-corrected bootstrap samples (with PROCESS v4.2, retina as a moderator, because PSD is dichotomous), and finally, cognition×group interaction terms were tested in parallel linear models.

All analyses were conducted in SPSS v.29.0.2 and R v4.4.1. Statistical significance was set at two-tailed p<.05. Analyses were completed between June 2024 and January 2025.

## 3. Results

Initially, 23,397 individuals met all quality control, inclusion, and exclusion criteria. Of these, 238 individuals with PSD (476 eyes) were identified and age- and sex-matched to 238 HC participants (476 eyes) (***Table 1***). The mean (SD) age across both groups was 53.5(8.4) years, with 47.1% identified as male. Most participants identified as White (87.4% PSD; 91.6% HC). A greater proportion of PSD individuals reported very low annual household income (<£18,000; *p*<.001) and a higher mean TDI score (*p*<.001) compared to HC.

**Table 1.**
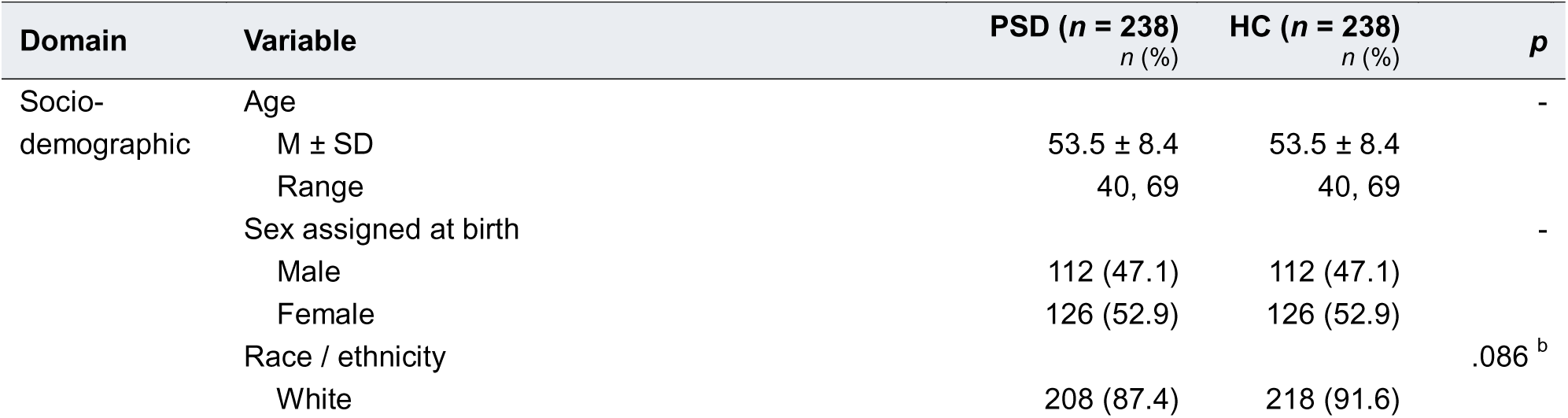
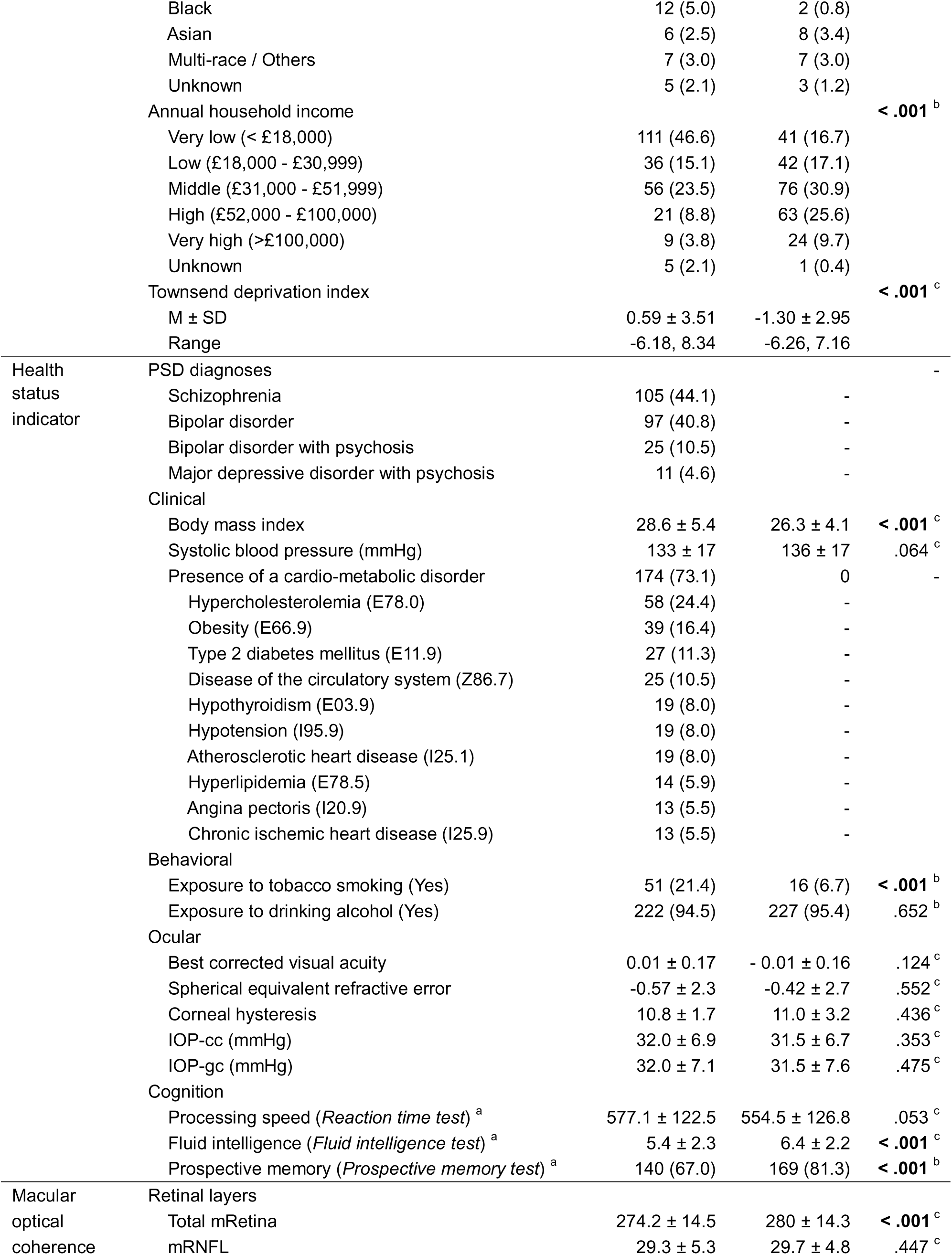
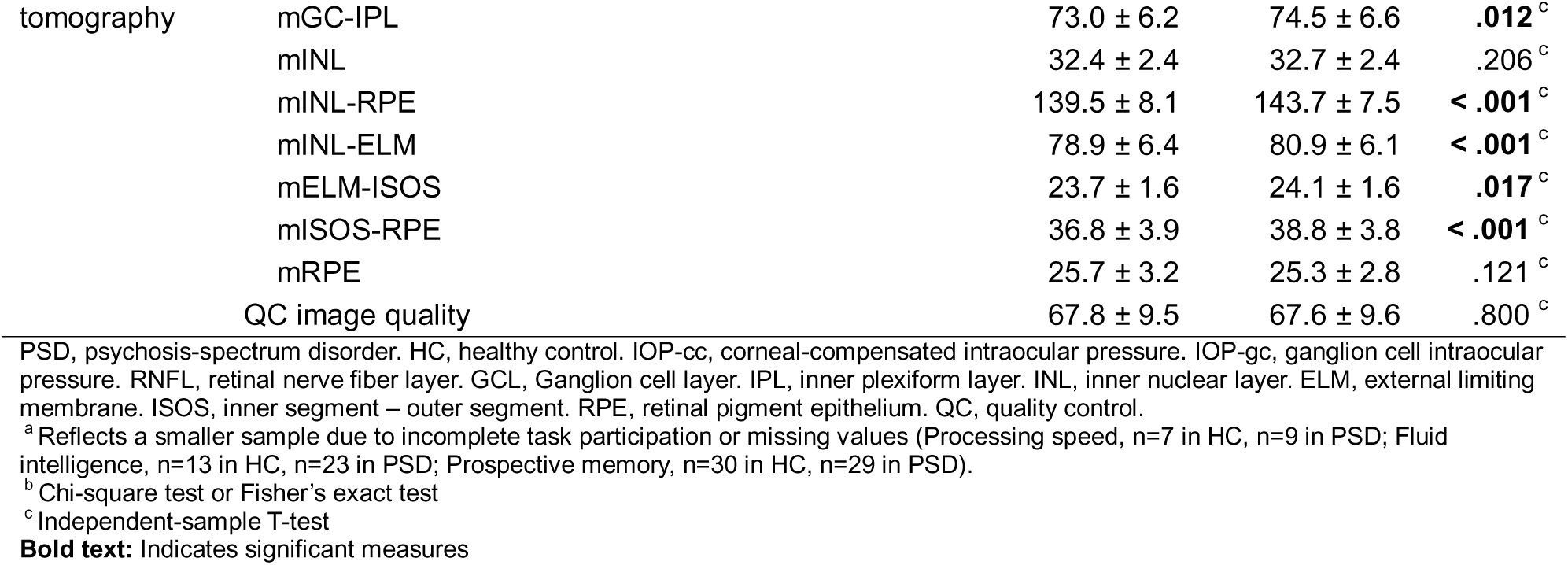
Cohort characteristics, by group.

Within PSD, diagnoses included SZ (44.1%), BD (44.5%), BD-P (6.7%), and MDD-P (4.6%). PSD participants had significantly higher BMI (*p*<.001) but no difference in systolic blood pressure compared to HC. Cardiometabolic disorders were present in 73.1% of the PSD group. Tobacco smoking exposure was more common in PSD (*p*<.001), while alcohol use did not differ between the groups. No significant group differences were observed in ocular measures, including BCVA, corneal hysteresis, IOP-cc, or IOP-gc (all *p*>.10). Cognitive performance was lower in the PSD group, with lower fluid intelligence (*p*<.001) and poorer prospective memory performance (*p*<.001).

Macular OCT measures indicated thinner mRetina (*p*<.001), mGC-IPL (*p*=.012), mINL-RPE (*p*<.001), mINL-ELM (*p*<.001), mELM-ISOS (*p*=.016), and mISOS-RPE (*p*<.001) in the PSD group. The mRNFL, mINL, and mRPE thicknesses did not differ significantly in the PSD group (*p*>.10) (***Table 1***).

### Identification of covariates influencing retinal thickness

To identify confounders of the association between PSD and retinal structure, linear regression models adjusted for age and sex demonstrated that greater TDI, BMI, and BCVA (poorer vision) scores were each associated with lower thickness in two or more retinal layers, while lower corneal hysteresis was associated with lower thickness in three layers (***Supplementary Tables 1–9***). Ethnicity was also associated with mRetina thickness. White participants had greater mRetina thickness (β=0.20, B=9.69, 95%CI [5.44, 13.95], *p*<.001), whereas Black participants had lower mRetina thickness (β=-0.19, B=-16.30, 95%CI [−24.06, −8.54], *p*<.001). Additionally, very low annual household income was associated with smaller mRetina thickness (β=-0.10, B=-3.36, 95%CI [−6.28, −0.44], *p*=.024). Because both Black ethnicity and very low income were associated with higher TDI scores (Black: β=0.18, B=3.50, 95%CI [1.73, 5.27], *p*<.001; Very low income: β=0.35, B=2.56, 95%CI [1.93, 3.19], *p*<.001), mediation analyses were conducted.

Results showed that TDI partially mediated the association between ethnicity and mRetina thickness (B=-2.22, SE=0.85, *p*=.026), and significantly mediated the association between very low income and mRetina thickness (B=-2.59, SE=0.58, *p*=.010). Given these findings, TDI was included as a covariate in subsequent models in place of ethnicity or income (***Supplementary Results***).

### Case-control differences in macular OCT measurements

In Model 1, adjusting for age and sex resulted in similar retinal layer group differences to the univariate analysis between PSD and HCs. In Model 2, adjusting for age, sex, TDI, BMI, BCVA, spherical equivalent refractive error, and corneal hysteresis, PSD individuals demonstrated similar thinning results to Model 1, except for the mELM-ISOS and mGC-IPL layers, which were no longer significant. Compared with HCs, PSD individuals without cardiometabolic disorders had thinner mRetina and mINL-ELM; however, only mINL-ELM remained significant after correction for multiple comparisons (***Table 2***). The effect sizes for significant case-control differences were predominantly small in all Models. However, in Model 1, mINL-RPE and mISOS-RPE exhibited a medium effect size.

**Table 2.**
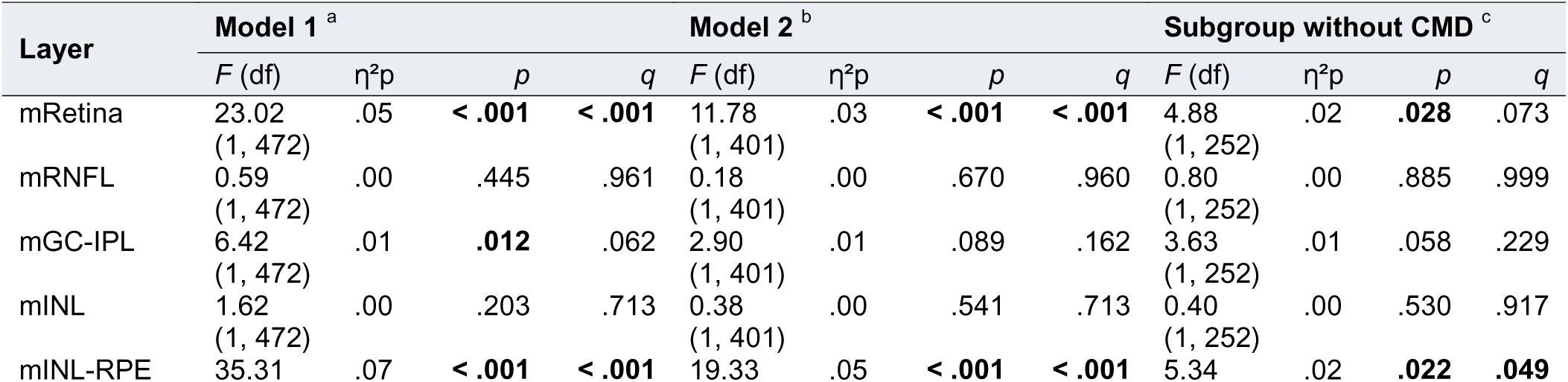
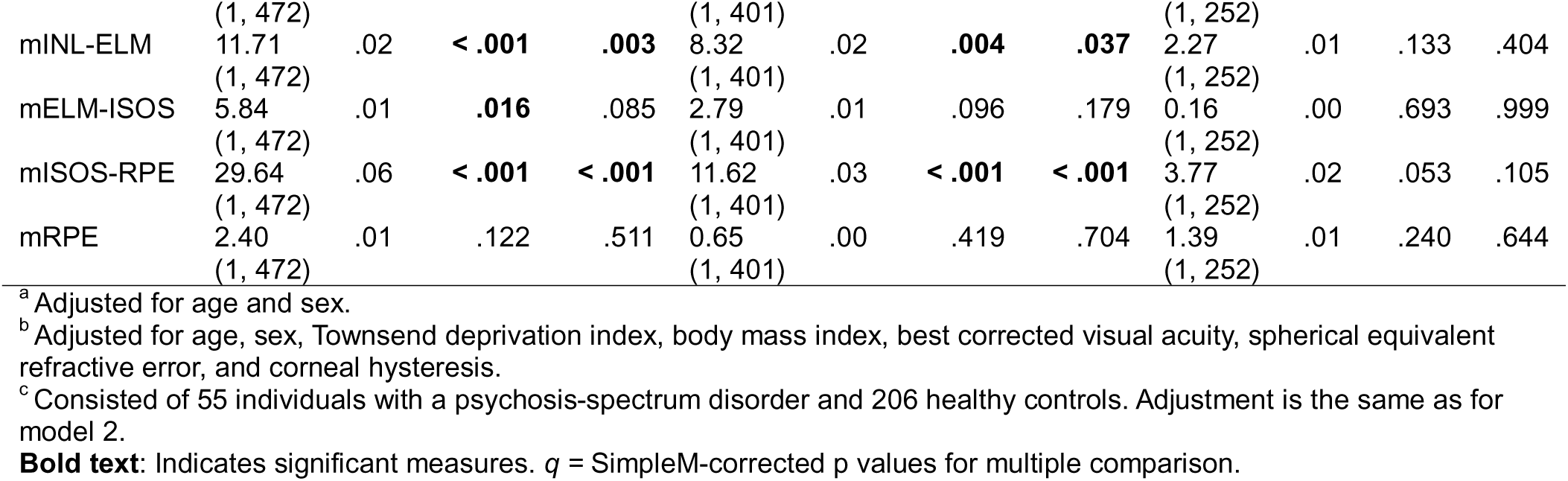
Case-control differences in macular OCT measurements, repeated-measures ANCOVA.

Subfield analyses were conducted for the photoreceptor layers (mINL-RPE, mINL-ELM, mELM-ISOS, mISOS-RPE) using Model 1 and 2 covariates (***Figure 2, see Supplementary Table 10-11*** *for subfield-level group comparisons*). Data demonstrated smaller mINL-ELM thickness in PSD was most pronounced in the central (F=30.93, η²p=.06, p<.001), followed by the inner (F=20.07, η²p=.04, p<.001) and outer (F=7.25, η²p=.02, p=.016) subfields. Conversely, smaller mISOS-RPE thickness in PSD was most pronounced in the outer (F=29.81, η²p=.06, p<.001), followed by the inner (F=23.49, η²p=05, p<.001) and central (F=19.39, η²p=04, p<.001) subfields.

**Figure 2.**
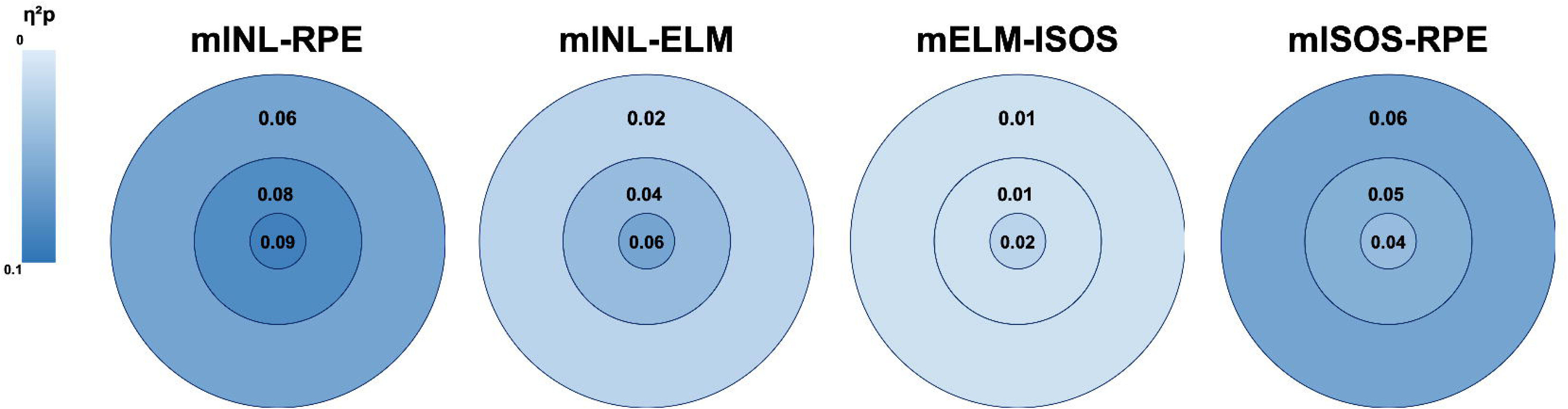
Diagrams showing subfield group differences, in microns, in photoreceptor layers between individuals with psychosis-spectrum disorders and healthy controls. Partial eta-squared (η^2^p) effect sizes from Model 1 (adjusted for age and sex) are displayed in each subfield, with color intensity indicating effect size.

Across diagnostic groups, mINL-RPE was the thinnest in SZ (*F*=26.84, η²p=.07, *p*<.001), followed by BD (*F*=16.60, η²p=.05, *p*=.006) and BD-P (*F*=12.37, η²p=.05, *p*<.001) compared to HC (see ***Supplementary Table 12-15*** *for comparisons of specific diagnostic subgroups*). MDD-P showed no significant mINL-RPE differences, though this group included few participants (*n*=11).

### Associations between retinal thickness and cognition

After adjusting for age and sex, lower thickness in the mINL-RPE and mINL-ELM layers was associated with poorer performance on the prospective memory test (ß=0.12, B=2.15, 95%CI [0.39, 3.92], p=.017 and ß=0.10, B=1.42, 95%CI [0.02, 2.83], p=.047, respectively), which was mediated by PSD status. The Sobel test showed significant indirect effects for both layers, point effect=0.76 (*p*=.004) for mINL-RPE and 0.35 (*p*=.025) for mINL-ELM (***Figure 3, Supplementary Table 16***). Bootstrap-based indirect effects were also significant for both layers (mINL-RPE ab=-0.17, 95%BC-CI [−0.30, −0.05]; mINL-ELM ab=-0.08, 95%BC-CI [−0.17, −0.01]. We additionally fitted prospective memory×diagnostic-group interaction models; the interaction term was non-significant for mINL-RPE (F=0.013, p=0.91) and mINL-ELM (F=0.343, p=0.558). Processing speed was also positively associated with RPE thickness, and fluid intelligence was positively associated with mRNFL and mELM-ISOS (***Supplementary Table 16***).

**Figure 3.**
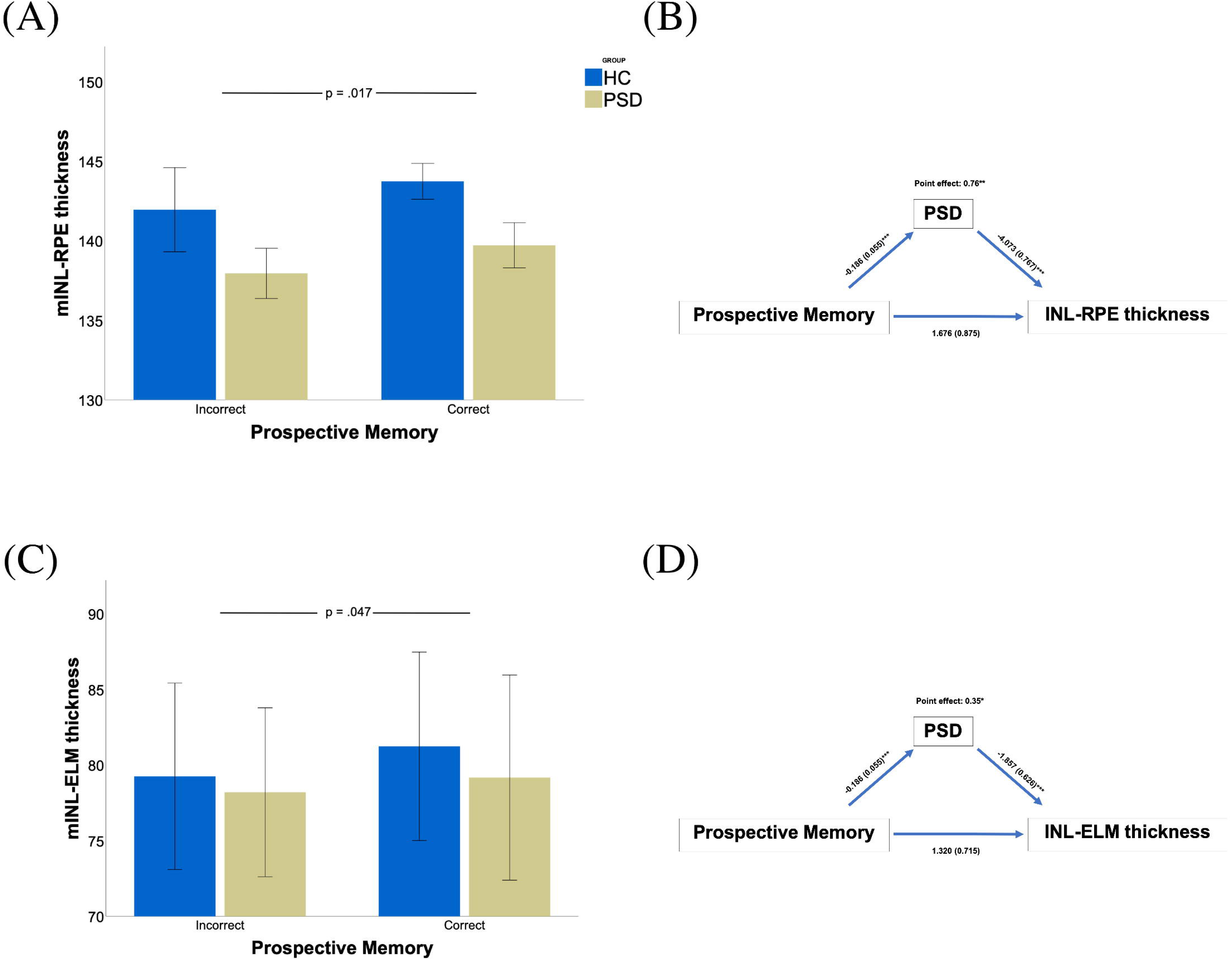
Associations between photoreceptor layers and prospective memory. ***3A.*** Lower mINL-RPE thickness was linked to poorer prospective memory performance, adjusting for age and sex (ß=0.12, B=2.15, 95% CI [0.39, 3.92], *p*=.017). ***3B.*** Incorrect answer in the prospective memory test was significantly associated with the PSD group (ß=-0.16, B=-0.19, 95% CI [−0.29, −0.08], *p*<.001). Prospective memory was not significantly associated with mINL-RPE thickness when adjusting for the PSD status (ß=0.09, B=1.68, 95% CI [-0.04, 3.40], *p*=.056). PSD was significantly associated with INL-RPE thinning when adjusting for prospective memory (ß=-0.25, B=-4.07, 95% CI [−5.58, −2.57], *p*<.001). The indirect effect estimate was 0.76 between the correct answer in the prospective memory task and INL-RPE thickness through the classification of PSD at the p-value of 0.004 (B=2.85, SE=0.27), as shown in the Sobel Test. ***3C.*** Lower mINL-ELM thickness was linked to poorer prospective memory performance, adjusting for age and sex (ß=0.10, B=1.42, 95% CI [0.02, 2.83], *p*=.047). ***3D.*** Prospective memory was not significantly associated with mINL-ELM thickness when adjusting for the PSD group (ß=0.09, B=1.32, 95% CI [-0.09, 2.73], *p*=.065). PSD was significantly associated with INL-ELM thinning when adjusting for prospective memory (ß=-0.15, B=-1.86, 95% CI [−3.09, −0.63], *p*=.003). The indirect effect estimate was 0.35 between the correct answer in prospective memory and INL-ELM thickness through the classification of PSD at the *p*-value of 0.025 (B=2.23, SE=0.15), as shown in the Sobel Test.

## 4. Discussion

Among 476 individuals (952 eyes) with OCT imaging that met quality control, inclusion, and exclusion criteria, we found that individuals with PSD had lower thickness of the mRetina, mGC-IPL (trending after correction), mINL-RPE, mINL-ELM, mELM-ISOS, and mISOS-RPE compared to HCs while adjusting for age and sex. After accounting for TDI, BMI, BCVA, spherical equivalent refractive error, and corneal hysteresis, these findings extended primarily to the photoreceptor layers, including thinner mINL-RPE, mINL-ELM, and mISOS-RPE thickness in the PSD compared to HCs. With cardiometabolic disorders excluded from the PSD group, and after covariate adjustment and multiple-comparison correction, only the mINL–RPE layer remained significantly thinner in PSD compared with HCs. Subfield analysis demonstrated that the mINL-ELM layer was most impacted from central>inner>outer regions, while the mISOS-RPE layer was most impacted from outer>inner>central regions. The SZ and BD groups demonstrated overlapping alterations in the mINL-RPE layer. Lastly, smaller photoreceptor layer thickness was associated with poorer prospective memory performance and lower fluid intelligence. To the best of our knowledge, this report is the largest and most comprehensive analysis of retinal layers across the psychosis spectrum, which supports evidence of photoreceptor dysfunction that is related to cognitive dysfunction in PSD.

### Inner layer associations with PSD

This study identified lower mGC-IPL thickness in individuals with PSD, though with a small effect size and trend-level after correction, and no significant differences in mRNFL or mINL thickness, consistent with prior meta-analyses [18,22,53]. However, mGC-IPL thickness did not remain statistically significant after adjusting for confounders, including ocular parameters such as BCVA and refractive error, both of which were associated with this layer (see ***Supplemental Table 3***). These findings highlight the need for OCT studies to routinely control for ocular parameters, particularly BCVA and refractive error. Smaller mGC-IPL thickness has also been observed in unaffected siblings of individuals with SZ or BD, suggesting a possible endophenotypic effect [54–56]. It has also been linked to greater polygenic risk in SZ, whereas mRNFL and mINL have not shown similar associations [33]. Supporting this, Boudriot et al. [6,34] found that synaptic-pathway genetic variants mediate the relationship between GC-IPL thinning, brain structure, and cognitive function, reinforcing the hypothesis that synaptic dysfunction contributes to both retinal and cortical alterations in PSD. Although genetic analysis of retinal cell types in neuropsychiatric disorders identified an enrichment of SZ genes with the expression profiles of amacrine cells, horizontal cells, and synapse biology pathways [34], the mINL—which comprises bipolar, amacrine, and horizontal cell bodies—remained preserved in our sample, suggesting selective involvement of retinal layers. In our study, mGC-IPL thickness was influenced by ethnicity and ocular factors but not by social determinants such as income level or TDI, pointing toward a heritable contribution, supported by an estimated mGC-IPL heritability of ∼46% [11]. A recent UKB study by Rabe et al. [57] further supports this view, reporting thinner mGC-IPL in individuals with higher SZ polygenic risk, particularly within neuroinflammatory pathways. Together, these findings suggest ganglion cell degeneration and/or reduced synaptic connectivity in PSD, likely shaped by genetic, ocular, and cardiometabolic factors, though further mechanistic research is warranted.

### Outer layer associations with PSD

Although studies of the outer retinal layers are limited, lower ONL thickness has been reported in chronic SZ and BD [14,18,58], but not in clinical high-risk or first-episode psychosis [33]. One study observed reduced photoreceptor inner segment thickness in SZ, though cone outer segment tips were preserved [58]. ONL thinning has been associated with poorer clinical outcomes, including greater negative symptoms and cognitive impairment, as well as with increased brain atrophy—such as reduced cortical thickness and visual network dysconnectivity—in both chronic psychosis and the general population [11,14,58]. Despite these findings, no comprehensive investigation of the inner and outer segments of the photoreceptor had been conducted in PSD. In our study, reduced mINL-RPE thickness was observed in PSD after adjusting for demographic, ocular, social, and cardiometabolic factors and correcting for multiple comparisons. mINL–RPE thickness was associated with SZ and BD-P, but not with BD or MDD-P (see ***Supplemental Tables 12–15***), suggesting that SZ and BD-P may share common retinal structural alterations. The effect size differences were more pronounced in older participants (see ***Supplemental Tables 17–18*** *for age-stratified results*), supporting the possibility that structural retinal changes emerge later in life, consistent with a neurodegenerative trajectory [31]. Photoreceptor heritability varies across layers: mINL-RPE and mINL-ELM show high heritability (∼67% in both), while mELM-ISOS (∼24%) and mISOS-RPE (∼33%) show lower heritability [11], indicating that some of these layers may serve as tractable phenotypes for future genomic studies exploring shared genetic architecture.

Meanwhile, electroretinography studies have shown cone and rod waveform alterations in unaffected youth at risk for psychosis [35–37], suggesting that functional impairments may arise earlier in life. This supports models in which retinal functional changes precede structural degeneration, potentially reflecting the cumulative effects of anterograde, retrograde, or shared developmental mechanisms, also modulated by cardiometabolic and ocular factors.

Subfield analyses revealed distinct morphological patterns: mINL-ELM (OPL and ONL) thinning was greatest in the central subfield, while mISOS-RPE (photoreceptor outer segment) thinning was most pronounced peripherally. As the central retina is cone-dominated and the peripheral retina is rod-dominated [59] (Figure 1C), these changes may translate into alterations in high-acuity/color vision under bright conditions and motion/night vision in dark environments [60]. Together, these findings suggest photoreceptor degeneration in PSD, particularly affecting dendrites, cell bodies, and outer segments. Further investigation into gene–environment interactions and underlying mechanisms is warranted to clarify the role of retinal alterations in the pathophysiology of PSD.

### Retina-cognition associations

In this study, smaller thickness in the mINL-RPE and mINL-ELM layers was associated with poorer prospective memory, while decreased mRNFL and mELM-ISOS thickness was linked to lower fluid intelligence, supporting prior evidence that retinal morphology is associated with cognition [6,40,61,62]. Photoreceptors contribute to cognition by providing sensory input for perception, learning, attention, and emotion; their disruption may contribute to cognitive deficits, especially in attention, recognition, and information processing [60,63]. Additionally, retinal axons send visual information to the brain and may influence reasoning and adaptive behavior [64], suggesting that their structural changes could be related to both visual deficits and lower fluid intelligence in people with PSD. The diagnosis × prospective memory interactions suggest that the association between photoreceptor thickness and prospective memory is comparable across groups (not moderated by diagnostic status); however, interaction tests are often underpowered and our subsamples were reduced, so small moderation effects cannot be ruled out. Longitudinal studies beginning in childhood and continuing into late-adulthood are needed to clarify causality, underlying mechanisms, and whether retina-cognition associations function as biomarkers of both neurodevelopmental vulnerability and later neuroprogression in PSD [31,65].

## Limitations

Although this is a mid-large, population-based, nested case–control (1:1 design) study utilizing data from the UKB, the findings may not be fully generalizable due to a few limitations. First, the sample is predominantly White and restricted to ages 40-69; volunteer bias and age-related retinal or cognitive decline may confound disease-specific effects despite statistical adjustment [66]. Second, the cross-sectional design of this study limits the ability to establish causation or examine the longitudinal trajectories of retinal layer changes, since the changes observed here may represent downstream effects of primary brain pathology (anterograde), retrograde trans-synaptic degeneration, or shared developmental mechanisms that affect both eye and brain, as well as cognitive changes. Additionally, Sobel mediation tests rely on an assumption of normally distributed indirect effects; given the cross-sectional design, the mediation findings should be viewed as exploratory. Future studies should include younger populations and encompass various stages of psychosis to enhance generalizability [67,68]. Third, while we conducted additional analyses to account for cardiometabolic disorders, the potential impact of other possible confounders, such as patients’ medication status [69] and residual confounding from unmeasured comorbidities on retinal outcomes, should be considered. The UKB lacks detailed clinical data (symptom scales and medication history) and some cognitive measures (Matrices test or paired association learning test) had a high level of missingness, so we could not relate findings to some clinical/cognitive variables. Furthermore, psychosis cases in UKB are higher-functioning and less severely affected than clinic cohorts, likely attenuating our effect sizes and limiting generalizability [70]. Fourth, the cognitive tasks available in UKB are abridged, single-trial, or short-form measures that correlate only moderately with gold-standard neuropsychological tests [50]. Future studies might use standard batteries such as MATRICS. Lastly, the absence of a non-psychotic psychiatric control group limits our ability to determine whether retinal and cognitive associations are specific to psychosis or reflect broader transdiagnostic effects.

## Conclusion

This study advances the understanding of retinal alterations in PSD by demonstrating smaller thickness in the mRetina, mINL-RPE, mINL-ELM, and mISOS-RPE layers, and a trend toward thinner mGC-IPL, after adjusting for confounders. These findings build on prior work by identifying outer layer involvement, specifically in the photoreceptors. Alterations in both ganglion cell–at trend level– and photoreceptor layers suggest disruptions across multiple levels of visual processing and highlight the potential of retinal measures as noninvasive markers of underlying genetic, ocular, cardiometabolic, environmental, and pathophysiological processes in PSD. Photoreceptor thickness was also associated with cognitive performance – particularly prospective memory and fluid intelligence – supporting the hypothesis that retinal alterations may reflect broader neural dysfunction in PSD. Longitudinal studies are needed to determine whether retinal alterations precede illness onset, track clinical progression, and/or are biomarkers of neurodegeneration in PSD. Clarifying these associations may support the development of retinal biomarkers for early detection, monitoring, and staging throughout disease progression.

## Data Availability

All data produced are available at UK Biobank

## Acknowledgments

This research has been conducted using the UKB Resource under application number 75692. We gratefully acknowledge the UKB for providing open access to this invaluable resource, which supports student researchers and advances health research. Furthermore, this work uses data provided by patients and collected by the NHS as part of their care and support. We sincerely thank the participants who generously contributed their health data, as the UKB relies on the dedication of volunteers committed to fostering health research and promoting a healthier society. Lastly, we recognize the Biomedical Sciences Career Program at Harvard Medical School for facilitating collaboration among the authors.

Supplementary information is available at MP’s website.

