## Supplement for "Retinal Cytoarchitectural Alterations Across the Psychosis Spectrum and Their Correlates with Cognition: A UK Biobank Nested Case-Control Study"

**Results**

***Linear regressions on retinal layer thickness and other variables***

***Supplementary Table 1*** presents factors associated with total macular thickness (mRetina). Linear regression analyses adjusted for age and sex indicated that race, very low income, higher TDI, BCVA, and corneal hysteresis were independently associated with mRetina thickness. Specifically, White participants had greater macular thickness (*p* < .001), whereas Black participants (*p* < .001), those with very low income (*p* = .024), and those with higher TDI scores (*p* < .001) exhibited significantly thinner macula. Additionally, worse BCVA (*p* = .004) and higher corneal hysteresis (*p*=.010) were associated with increased macular thickness.

***Supplementary Table 2*** summarizes variables significantly associated with mRNFL thickness. After adjusting for age and sex, the White race (*p*= 042), the Black race (*p*=.040), and worse BCVA (*p*<.001) were independently associated with mRNFL thickness. Specifically, White participants had greater mRNFL thickness, whereas Black participants and those with poorer visual acuity exhibited lower thickness.

***Supplementary Table 3*** describes the factors associated with mGC-IPL thickness. Linear regression analyses (adjusted by age and sex) revealed that being White (*p*=.031) and having lower BCVA (*p*=.015) were associated with increased mGC-IPL thickness.

***Supplementary Table 4*** reports variables associated with mINL thickness (bipolar cells). After adjusting for age and sex, Black race was the only factor significantly associated with lower mINL thickness (*p*=.007), indicating thinner bipolar cell layers in this group than others.

***Supplementary Table 5*** describes the factors associated with mINL-RPE (photoreceptor layer) thickness. Linear regression analyses (adjusted by age and sex) revealed that race, income, TDI, prospective memory, and corneal hysteresis were associated with mINL-RPE thickness. Specifically, while being White (*p*<.001), having high income (*p*=.005), higher prospective memory score (*p*<.017), and corneal hysteresis (*p*=.007) were associated with increased thickness, being Black (*p*<.001) or multi-race/others (*p*=0.033), having very low income (*p*<0.001), greater TDI (*p*<.001) were associated with decreased mINL-RPE thickness.

***Supplementary Table 6*** summarizes the factors associated with mINL-ELM (OPL and ONL) thickness. Linear regression analyses (adjusted by age and sex) revealed that race, income, TDI, prospective memory, and corneal hysteresis were associated with mINL-ELM thickness. Specifically, while being White (*p*<.001), higher prospective memory score (*p*<.017), and greater corneal hysteresis (*p*=.007) were associated with increased thickness, being Black (*p*<.001) or Asian (*p*=.027), having very low income (*p*<0.03), greater TDI (*p*=.004) were associated with decreased mINL-ELM thickness.

***Supplementary Table 7*** presents the variables associated with mELM-ISOS (photoreceptor inner segment) thickness. Linear regression analyses (adjusted by age and sex) revealed that race, income, TDI, BMI, and fluid intelligence were associated with mELM-ISOS thickness. Specifically, being Asian (*p*=.012), having very high income (p=.003), and reporting higher fluid intelligence (*p*=.001) were associated with increased thickness, being multi-race/others (*p*=.021), having very low income (*p*=.013), greater TDI (*p*=.001), greater BMI (*p*=.001) were associated with decreased ELM-ISOS thickness.

***Supplementary Table 8*** reports the factors associated with mISOS-RPE (photoreceptor outer segment) thickness. Linear regression analyses (adjusted by age and sex) revealed that race, income, TDI, BMI, and BCVA were associated with ISOS-RPE thickness. Specifically, while being White (*p*=.018) and having high income (*p*=.006) were associated with increased thickness, being Black (*p*<.001), having very low income (*p*<.001), greater TDI (*p*=.002), greater BMI (*p*=.001), and greater BCVA (*p*=.042) were associated with decreased mISOS-RPE thickness.

***Supplementary Table 9*** summarizes variables associated with mRPE thickness. After adjusting for age and sex, Black race (*p*<.001), White race (*p*=.002), very low income (p=.012), higher TDI (*p*=.003), and higher systolic blood pressure (*p*=.039) were significantly associated with mRPE thickness. Black participants had greater mRPE thickness, while White participants showed lower thickness. Thicker mRPE was also observed in individuals with very low income and those with higher TDI or systolic blood pressure

**Levene's Test of Equality of Error Variances for Retinal Layers** (left / right eye, respectively): mRetina (p=0.478 / p=0.914), mRNFL (p=0.099 / p=0.771), mGC-IPL (p=0.563 / p=0.406), mINL (p=0.569 / p=0.870), mINL-RPE (p=0.633 / p=0.353), mINL-ELM (p=0.794 / p=0.240), mELM-ISOS (p=0.115 / p=0.176), mISOS-RPE (p=0.953 / p=0.389), mRPE (p=0.497 / p=0.561)

| **Supplementary Table 1.** Linear regression analyses identifying variables associated with Total mRetina layer thickness | | | | |
| --- | --- | --- | --- | --- |
| **Variable** | **β** | **B** | **95% CI** | ***p*** ^†^ |
| Race / ethnicity |  |  |  |  |
| White | 0.20 | 9.69 | (5.44, 13.95) | **< .001** |
| Black | -0.19 | -16.30 | (-24.06, -8.54) | **< .001** |
| Asian | -0.13 | -1.16 | (-8.99, 6.68) | .771 |
| Multi-race / Others | -0.10 | -8.48 | (-16.31, -0.65) | **.034** |
| Unknown | -0.09 | -10.33 | (-20.60, -0.07) | **.049** |
| Income |  |  |  |  |
| Very low (< £18,000) | -0.10 | -3.36 | (-6.28, -0.44) | **.024** |
| Low (£18,000 - £30,999) | -0.03 | -1.17 | (-4.74, 2.41) | .522 |
| Middle (£31,000 - £51,999) | 0.05 | 1.51 | (-1.41, 4.43) | .310 |
| High (£52,000 - £100,000) | 0.09 | 3.44 | (-0.16, 7.04) | .061 |
| Very high (>£100,000) | 0.08 | 4.33 | (-0.80, 9.46) | .098 |
| Unknown | -0.13 | -17.35 | (-29.16, -5.53) | **.004** |
| Other variables |  |  |  |  |
| Townsend deprivation index | -0.15 | -0.67 | (-1.06, -0.28) | **< .001** |
| Body mass index | -0.08 | -0.24 | (-0.52, 0.03) | .081 |
| Systolic blood pressure | -0.02 | -0.02 | (-0.10, 0.06) | .636 |
| Exposure to tobacco smoking | -0.03 | -1.23 | (-5.08, 2.61) | .530 |
| Exposure to drinking alcohol | 0.01 | 0.41 | (-5.65, 6.48) | .893 |
| Visual acuity | -0.14 | -11.91 | (-19.92, -3.90) | **.004** |
| Spherical equivalent refractive error | 0.29 | 0.05 | (0.03, 0.07) | **< .001** |
| Corneal hysteresis | 0.13 | 0.71 | (0.17, 1.26) | **.010** |
| IOP-cc (mmHg) | -0.07 | -0.15 | (-0.35, 0.05) | .130 |
| IOP-gc (mmHg) | -0.01 | -0.01 | (-0.20, 0.17) | .912 |

^†^ Adjusted by age and gender.

| **Supplementary Table 2.** Linear regression analyses identifying variables associated with mRNFL | | | | |
| --- | --- | --- | --- | --- |
| **Variable** | **β** | **B** | **95% CI** | ***p*** ^†^ |
| Race / ethnicity |  |  |  |  |
| White | 0.09 | 1.54 | (0.05, 3.03) | **.042** |
| Black | -0.09 | -2.83 | (-5.54, -0.12) | **.040** |
| Asian | -0.02 | -0.59 | (-3.29, 2.11) | .667 |
| Multi-race / Others | -0.47 | -1.42 | (-4.12, 1.29) | .303 |
| Unknown | -0.10 | -0.41 | (3.96, 3.14) | .820 |
| Income |  |  |  |  |
| Very low (< £18,000) | -0.06 | -0.62 | (-1.63, 0.38) | .225 |
| Low (£18,000 - £30,999) | -0.03 | -0.42 | (-1.65, 0.81) | .502 |
| Middle (£31,000 - £51,999) | -0.01 | -0.01 | (-1.02, 1.00) | .986 |
| High (£52,000 - £100,000) | 0.05 | 0.68 | (-0.56, 1.92) | .283 |
| Very high (>£100,000) | 0.07 | 1.43 | (-0.34, 3.19) | .114 |
| Unknown | 0.01 | 0.09 | (-4.02, 4.19) | .967 |
| Other variables |  |  |  |  |
| Townsend deprivation index | -0.05 | -0.07 | (-0.21, 0.07) | .307 |
| Body mass index | -0.08 | -0.08 | (-0.17, 0.16) | .102 |
| Systolic blood pressure | -0.06 | -0.02 | (-0.05, 0.01) | .230 |
| Exposure to tobacco smoking | -0.04 | -0.59 | (-1.91, 0.74) | .385 |
| Exposure to drinking alcohol | 0.02 | 0.45 | (-1.64, 2.53) | .673 |
| Best corrected visual acuity | -0.15 | -4.70 | (-7.46, -1.94) | **< .001** |
| Spherical equivalent refractive error | -0.14 | -0.07 | (-0.13, -0.02) | **.004** |
| Corneal hysteresis | 0.06 | 0.11 | (-0.06, 0.28) | .209 |
| IOP-cc (mmHg) | 0.02 | 0.01 | (-0.06, 0.08) | .739 |
| IOP-gc (mmHg) | 0.03 | 0.03 | (-0.04, 0.08) | .533 |

^†^ Adjusted by age and gender.

| **Supplementary Table 3.** Linear regression analyses identifying variables associated with mGC-IPL thickness (ganglion cells-bipolar cells) | | | | |
| --- | --- | --- | --- | --- |
| **Variable** | **β** | **B** | **95% CI** | ***p*** ^†^ |
| Race / ethnicity |  |  |  |  |
| White | 0.10 | 2.08 | (0.19, 3.97) | **.031** |
| Black | -0.05 | -1.93 | (-5.38, 1.52) | .272 |
| Asian | 0.02 | 0.75 | (-2.68, 4.17) | .668 |
| Multi-race / Others | -0.06 | -2.35 | (-5.78, 1.08) | .179 |
| Unknown | -0.11 | -5.70 | (-10.18, -1.23) | **.013** |
| Income |  |  |  |  |
| Very low (< £18,000) | 0.03 | 0.38 | (-0.90, 1.66) | .557 |
| Low (£18,000 - £30,999) | -0.07 | -1.14 | (-2.70, 0.42) | .150 |
| Middle (£31,000 - £51,999) | 0.04 | 0.51 | (-0.77, 1.78) | .437 |
| High (£52,000 - £100,000) | -0.01 | -0.03 | (-1.61, 1.55) | .969 |
| Very high (>£100,000) | 0.04 | 0.96 | (-1.28, 3.21) | .400 |
| Unknown | -0.12 | -6.89 | (-12.06, -1.72) | **.009** |
| Other variables |  |  |  |  |
| Townsend deprivation index | -0.04 | -0.08 | (-0.26, 0.09) | .340 |
| Body mass index | -0.02 | -0.03 | (-0.15, 0.10) | .684 |
| Systolic blood pressure | -0.02 | -0.01 | (-0.04, 0.03) | .710 |
| Exposure to tobacco smoking | -0.03 | -0.49 | (-2.17, 1.20) | .571 |
| Exposure to drinking alcohol | -0.07 | -2.08 | (-4.72, 0.56) | .122 |
| Best corrected visual acuity | -0.11 | -4.36 | (-7.88, -0.85) | **.015** |
| Spherical equivalent refractive error | 0.32 | 0.13 | (0.10, 0.17) | **< .001** |
| Corneal hysteresis | 0.06 | 0.14 | (-0.09, 0.37) | .224 |
| IOP-cc (mmHg) | -0.06 | -0.06 | (-0.14, 0.03) | .205 |
| IOP-gc (mmHg) | -0.03 | -0.03 | (-0.11, 0.05) | .484 |

^†^ Adjusted by age and gender.

| **Supplementary Table 4.** Linear regression analyses identifying variables associated with mINL thickness (bipolar cells) | | | | |
| --- | --- | --- | --- | --- |
| **Variable** | **β** | **B** | **95% CI** | ***p*** ^†^ |
| Race / ethnicity |  |  |  |  |
| White | 0.09 | 0.68 | (-0.02, 1.38) | .058 |
| Black | -0.12 | -1.75 | (-3.02 -0.48) | **.007** |
| Asian | 0.02 | 0.27 | (-1.00, 1.54) | .671 |
| Multi-race / Others | -0.01 | -0.08 | (-1.35, 1.20) | .907 |
| Unknown | -0.06 | -1.19 | (-2.86, 0.48) | .162 |
| Income |  |  |  |  |
| Very low (< £18,000) | 0.01 | 0.01 | (-0.47, 0.48) | .991 |
| Low (£18,000 - £30,999) | 0.01 | 0.03 | (-0.55, 0.61) | .916 |
| Middle (£31,000 - £51,999) | 0.05 | 0.28 | (-0.20, 0.75) | .252 |
| High (£52,000 - £100,000) | -0.01 | -0.04 | (-0.62, 0.55) | .903 |
| Very high (>£100,000) | -0.06 | -0.54 | (-1.37, 0.29) | .202 |
| Unknown | -0.08 | -1.69 | (-3.61, 0.24) | .086 |
| Other variables |  |  |  |  |
| Townsend deprivation index | -0.04 | -0.03 | (-0.10, 0.03) | .335 |
| Body mass index | -0.01 | -0.01 | (-0.05, 0.04) | .791 |
| Systolic blood pressure | 0.03 | 0.01 | (-0.01, 0.02) | .488 |
| Exposure to tobacco smoking | -0.03 | -0.17 | (-0.79, 0.46) | .596 |
| Exposure to drinking alcohol | -0.05 | -0.49 | (-1.47, 0.49) | .328 |
| Best corrected visual acuity | -0.01 | -0.10 | (-1.41, 1.21) | .876 |
| Spherical equivalent refractive error | 0.42 | 0.44 | (0.35, 0.53) | **< .001** |
| Corneal hysteresis | 0.06 | 0.05 | (-0.03, 0.14) | .230 |
| IOP-cc (mmHg) | -0.01 | -0.01 | (-0.03, 0.03) | .964 |
| IOP-gc (mmHg) | 0.03 | 0.01 | (-0.02, 0.04) | .472 |

^†^ Adjusted by age and gender.

| **Supplementary Table 5.** Linear regression analyses identifying variables associated with mINL-RPE thickness (photoreceptor layer thickness) | | | | |
| --- | --- | --- | --- | --- |
| **Variable** | **β** | **B** | **95% CI** | ***p*** ^†^ |
| Race / ethnicity |  |  |  |  |
| White | 0.21 | 5.39 | (3.01, 7.70) | **< .001** |
| Black | -0.21 | -9.79 | (-13.99, -5.59) | **< .001** |
| Asian | -0.03 | -1.59 | (-5.85, 2.67) | .463 |
| Multi-race / Others | -0.10 | -4.63 | (-8.89, -0.38) | **.033** |
| Unknown | -0.05 | -3.03 | (-8.63, 2.57) | .288 |
| Income |  |  |  |  |
| Very low (< £18,000) | -0.18 | -3.12 | (-4.69, -1.55) | **< .001** |
| Low (£18,000 - £30,999) | 0.02 | 0.37 | (-1.58, 2.31) | .711 |
| Middle (£31,000 - £51,999) | 0.04 | 0.74 | (-0.85, 2.33) | .361 |
| High (£52,000 - £100,000) | 0.13 | 2.83 | (0.87, 4.77) | **.005** |
| Very high (>£100,000) | 0.08 | 2.48 | (-0.31, 5.27) | .081 |
| Unknown | -0.12 | -8.86 | (-15.29, -2.43) | **.007** |
| Other variables |  |  |  |  |
| Townsend deprivation index | -0.20 | -0.48 | (-0.70, -0.27) | **< .001** |
| Body mass index | -0.08 | -0.13 | (-0.28, 0.02) | .078 |
| Systolic blood pressure | -0.01 | 0.01 | (-0.05, 0.04) | .984 |
| Exposure to tobacco smoking | 0.01 | 0.01 | (-2.01, 2.10) | .993 |
| Exposure to drinking alcohol | 0.07 | 2.54 | (-0.74, 5.82) | .129 |
| Best corrected visual acuity | -0.06 | -2.75 | (-7.14, 1.64) | .219 |
| Spherical equivalent refractive error | 0.24 | 0.07 | (0.05, 0.10) | **< .001** |
| Corneal hysteresis | 0.13 | 0.41 | (0.11, 0.71) | **.007** |
| IOP-cc (mmHg) | -0.09 | -0.11 | (-0.22, -0.01) | **.050** |
| IOP-gc (mmHg) | -0.01 | -0.01 | (-0.11, 0.09) | .804 |

^†^ Adjusted by age and gender.

| **Supplementary Table 6.** Linear regression analyses identifying variables associated with mINL-ELM thickness | | | | |
| --- | --- | --- | --- | --- |
| **Variable** | **β** | **B** | **95% CI** | ***p*** ^†^ |
| Race / ethnicity |  |  |  |  |
| White | 0.19 | 3.90 | (2.08, 5.72) | **< .001** |
| Black | -0.12 | -4.47 | (-7.82, -1.13) | **.009** |
| Asian | -0.10 | -3.77 | (-7.09, -0.44) | **.027** |
| Multi-race / Others | -0.07 | -2.76 | (-6.11, 0.58) | .105 |
| Unknown | -0.06 | -3.12 | (-7.51, 1.28) | .164 |
| Income |  |  |  |  |
| Very low (< £18,000) | -0.10 | -1.38 | (-2.62, -0.13) | **.030** |
| Low (£18,000 - £30,999) | 0.03 | 0.42 | (-1.11, 1.95) | .589 |
| Middle (£31,000 - £51,999) | 0.02 | 0.31 | (-0.93, 1.56) | .621 |
| High (£52,000 - £100,000) | 0.07 | 1.18 | (-0.36, 2.72) | .133 |
| Very high (>£100,000) | 0.03 | 0.74 | (-1.45, 2.94) | .507 |
| Unknown | -0.07 | -3.95 | (-9.03, 1.12) | .127 |
| Other variables |  |  |  |  |
| Townsend deprivation index | -0.13 | -0.25 | (-0.42, -0.08) | **.004** |
| Body mass index | 0.03 | 0.04 | (-0.08, 0.15) | .546 |
| Systolic blood pressure | 0.02 | 0.01 | (-0.03, 0.04) | .609 |
| Exposure to tobacco smoking | 0.03 | 0.60 | (-1.04, 2.24) | .473 |
| Exposure to drinking alcohol | 0.04 | 1.17 | (-1.42, 3.75) | .377 |
| Best corrected visual acuity | 0.01 | 0.28 | (-3.17, 3.73) | .874 |
| Spherical equivalent refractive error | 0.32 | 0.12 | (0.09, 0.16) | **< .001** |
| Corneal hysteresis | 0.13 | 0.32 | (0.09, 0.56) | **.007** |
| IOP-cc (mmHg) | -0.09 | -0.08 | (-0.17, 0.01) | .066 |
| IOP-gc (mmHg) | -0.01 | -0.01 | (-0.09, 0.07) | .774 |

^†^ Adjusted by age and gender.

| **Supplementary Table 7.** Linear regression analyses identifying variables associated with ELM-ISOS thickness | | | | |
| --- | --- | --- | --- | --- |
| **Variable** | **β** | **B** | **95% CI** | ***p*** ^†^ |
| Race / ethnicity |  |  |  |  |
| White | 0.02 | 0.09 | (-0.38, 0.56) | .705 |
| Black | -0.08 | -0.76 | (-1.60, 0.09) | .080 |
| Asian | 0.11 | 1.07 | (0.24, 1.91) | **.012** |
| Multi-race / Others | -0.11 | -0.99 | (-1.83, -0.15) | **.021** |
| Unknown | 0.05 | 0.62 | (-0.49, 1.73) | .270 |
| Income |  |  |  |  |
| Very low (< £18,000) | -0.11 | -0.40 | (-0.71, -0.08) | **.013** |
| Low (£18,000 - £30,999) | -0.05 | -0.21 | (-0.59, 0.18) | .288 |
| Middle (£31,000 - £51,999) | 0.01 | 0.05 | (-0.27, 0.36) | .765 |
| High (£52,000 - £100,000) | 0.07 | 0.30 | (-0.09, 0.69) | .128 |
| Very high (>£100,000) | 0.14 | 0.84 | (0.29, 1.39) | **.003** |
| Unknown | 0.02 | 0.29 | (-0.99, 1.57) | .659 |
| Other variables |  |  |  |  |
| Townsend deprivation index | -0.15 | -0.07 | (-0.11, -0.03) | **.001** |
| Body mass index | -0.15 | -0.05 | (-0.08, -0.02) | **.001** |
| Systolic blood pressure | -0.04 | -0.01 | (-0.01, 0.01) | .390 |
| Exposure to tobacco smoking | -0.06 | -0.26 | (-0.67, 0.16) | .220 |
| Exposure to drinking alcohol | 0.03 | 0.23 | (-0.42, 0.88) | .483 |
| Best corrected visual acuity | -0.08 | -0.78 | (-1.65, 0.08) | .077 |
| Spherical equivalent refractive error | -0.22 | -0.34 | (-0.49, -0.20) | **< .001** |
| Corneal hysteresis | 0.03 | 0.02 | (-0.04, 0.08) | .542 |
| IOP-cc (mmHg) | -0.01 | 0.01 | (-0.02, 0.02) | .984 |
| IOP-gc (mmHg) | 0.02 | 0.01 | (-0.02, 0.02) | .754 |

^†^ Adjusted by age and gender.

| **Supplementary Table 8.** Linear regression analyses identifying variables associated with ISOS-RPE thickness | | | | |
| --- | --- | --- | --- | --- |
| **Variable** | **β** | **B** | **95% CI** | ***p*** ^†^ |
| Race / ethnicity |  |  |  |  |
| White | 0.11 | 1.40 | (0.24, 2.56) | **.018** |
| Black | -0.20 | -4.56 | (-6.64, -2.48) | **< .001** |
| Asian | 0.05 | 1.10 | (-1.00, 3.20) | .302 |
| Multi-race / Others | -0.04 | -0.88 | (-2.99, 1.23) | .413 |
| Unknown | -0.02 | -0.54 | (-3.30, 2.23) | .704 |
| Income |  |  |  |  |
| Very low (< £18,000) | -0.16 | -1.35 | (-2.13, -0.57) | **< .001** |
| Low (£18,000 - £30,999) | 0.02 | 0.16 | (-0.81, 1.11) | .752 |
| Middle (£31,000 - £51,999) | 0.04 | 0.38 | (-0.41, 1.16) | .345 |
| High (£52,000 - £100,000) | 0.13 | 1.35 | (0.39, 2.31) | **.006** |
| Very high (>£100,000) | 0.06 | 0.90 | (-0.48, 2.28) | .199 |
| Unknown | -0.15 | -5.19 | (-8.35, -2.03) | **.001** |
| Other variables |  |  |  |  |
| Townsend deprivation index | -0.14 | -0.17 | (-0.27, -0.06) | **.002** |
| Body mass index | -0.15 | -0.12 | (-0.20, -0.05) | **.001** |
| Systolic blood pressure | -0.02 | -0.01 | (-0.03, 0.02) | .611 |
| Exposure to tobacco smoking | -0.03 | -0.33 | (-1.37, 0.70) | .527 |
| Exposure to drinking alcohol | 0.06 | 1.14 | (-0.47, 2.76) | .166 |
| Best corrected visual acuity | -0.10 | -2.25 | (-4.41, -0.08) | **.042** |
| Spherical equivalent refractive error | 0.08 | 0.05 | (-0.01, 0.11) | .099 |
| Corneal hysteresis | 0.05 | 0.07 | (-0.08, 0.22) | .359 |
| IOP-cc (mmHg) | -0.05 | -0.03 | (-0.08, 0.03) | .303 |
| IOP-gc (mmHg) | -0.01 | -0.01 | (-0.05, 0.05) | .864 |

^†^ Adjusted by age and gender.

| **Supplementary Table 9.** Linear regression analyses identifying variables associated with mRPE thickness | | | | |
| --- | --- | --- | --- | --- |
| **Variable** | **β** | **B** | **95% CI** | ***p*** ^†^ |
| Race / ethnicity |  |  |  |  |
| White | -0.14 | -1.39 | (-2.27, -0.51) | **.002** |
| Black | 0.19 | 3.43 | (1.84, 5.03) | **< .001** |
| Asian | 0.02 | 0.28 | (-1.33, 1.89) | .730 |
| Multi-race / Others | 0.07 | 1.18 | (-0.43, 2.79) | .151 |
| Unknown | -0.02 | -0.52 | (-2.64, 1.59) | .627 |
| Income |  |  |  |  |
| Very low (< £18,000) | 0.12 | 0.77 | (0.17, 1.37) | **.012** |
| Low (£18,000 - £30,999) | 0.01 | 0.03 | (-0.70, 0.77) | .935 |
| Middle (£31,000 - £51,999) | -0.08 | -0.55 | (1.15, 0.05) | .072 |
| High (£52,000 - £100,000) | -0.08 | -0.67 | (-1.41, 0.07) | .075 |
| Very high (>£100,000) | 0.03 | 0.35 | (-0.71, 1.41) | .516 |
| Unknown | 0.06 | 1.55 | (-0.90, 3.99) | .214 |
| Other variables |  |  |  |  |
| Townsend deprivation index | 0.14 | 0.12 | (0.04, 0.20) | **.003** |
| Body mass index | 0.04 | 0.02 | (-0.03, 0.08) | .428 |
| Systolic blood pressure | 0.10 | 0.02 | (0.01, 0.03) | **.039** |
| Exposure to tobacco smoking | 0.49 | 0.28 | (-0.51, 1.07) | .491 |
| Exposure to drinking alcohol | -0.06 | -0.84 | (-2.01, 0.40) | .185 |
| Best corrected visual acuity | 0.04 | 0.63 | (-1.03, 2.30) | .455 |
| Spherical equivalent refractive error | 0.07 | 0.05 | (-0.02, 0.13) | .163 |
| Corneal hysteresis | -0.04 | -0.05 | (-0.17, 0.06) | .387 |
| IOP-cc (mmHg) | 0.06 | 0.03 | (-0.02, 0.07) | .236 |
| IOP-gc (mmHg) | 0.01 | 0.01 | (-0.04, 0.04) | .938 |

^†^ Adjusted by age and gender.

***Race and total mRetina thickness***

White individuals exhibited significantly greater total mRetina thickness compared to non-White individuals (β=0.20, B=9.69, 95% CI [5.44, 13.95], *p*<.001). Conversely, Black individuals demonstrated significantly lower total mRetina thickness compared to non-Black individuals (β=-0.19, B=-16.30, 95% CI [-24.06, -8.54], *p*<.001). In addition, Black individuals had significantly higher TDI scores than non-Black individuals (β=0.18, B=3.50, 95% CI [1.73, 5.27], *p*<.001). However, mediation analysis via the Sobel test indicated that TDI partly mediated the relationship between Black race and total mRetina thickness (B=-2.22, SE=0.85, *p*=0.026), with an indirect effect estimate of -1.89.

Point effect: -1.89 **

Townsend deprivation

3.50 (0.90) ***	-0.54 (0.20) **

Total mRetina thickness

Black race

-14.40 (3.99) ***

SOBEL test: B = -2.22, SE = 0.85, p = 0.026

***Income and total mRetina thickness***

Individuals with very low income exhibited significantly lower total mRetina thickness than those without very low income (β=-0.10, B=-3.36, 95% CI [-6.28, -0.44], *p*=.024). Additionally, very low-income individuals had significantly higher TDI scores (β=0.35, B=2.56, 95% CI [1.93, 3.19], *p*<.001). Mediation analysis via the Sobel test demonstrated that TDI significantly mediated the relationship between very low income and total mRetina thickness (B=-2.59, SE=0.58, *p*=.010), with an indirect effect estimate of -3.14. TDI serves as a mediator in the relationship between very low income and total macular retinal thickness. It is recommended that TDI be utilized as a covariate instead of very low income.

Point effect: -3.14

Townsend deprivation

2.56 (0.32) ***	-0.58 (0.21) **

Very low income

Total mRetina thickness

-1.87(1.57)

SOBEL test: B = -2.59, SE = 0.58, p = 0.010

***OCT measurements in photoreceptor layers (by subfields)***

***Supplementary Tables 10 and 11*** present case-control differences in photoreceptor-associated macular layers across central, inner, and outer subfields.

In Model 1 (adjusted for age and sex), all four layers—mINL-RPE, mINL-ELM, mELM-ISOS, and mISOS-RPE—showed significant thickness reductions across all subfields. For mINL-RPE, mINL-ELM, and mELM-ISOS, the effect sizes followed a consistent central > inner > outer pattern (e.g., mINL-RPE: η²p = .09 central, .08 inner, .06 outer; mINL-ELM: η²p = .06, .04, .02). In contrast, mISOS-RPE showed an opposite pattern, with effect sizes following an outer > inner > central gradient (η²p = .06 outer, .05 inner, .04 central).

In Model 2 (adjusted for age, sex, TDI, BMI, BCVA, and corneal hysteresis), mINL-RPE, mINL-ELM, and mISOS-RPE remained significant across all subfields. The central > inner > outer pattern persisted for mINL-RPE (η²p = .07, .06, .04) and mINL-ELM (η²p = .05, .04, .02). Again, mISOS-RPE showed an outer > inner > central pattern (η²p = .04, .03, .02). mELM-ISOS was only significant in the central subfield in Model 2.

| **Supplementary Table 10.** Case-control differences in macular OCT measurements (photoreceptor layers), by subfield, Model 1 | | | | | | | | | | | |
| --- | --- | --- | --- | --- | --- | --- | --- | --- | --- | --- | --- |
| **Layer** | **Central subfield** ^a^ | | |  | **Inner subfield** ^a^ | | |  | **Outer subfield** ^a^ | | |
|  | *F* (df) | η²p | *p* |  | *F* (df) | η²p | *p* |  | *F* (df) | η²p | *p* |
| mINL-RPE | 46.67 (1,472) | .09 | **< .001** |  | 39.82 (1,472) | .08 | **< .001** |  | 29.13 (1,472) | .06 | **< .001** |
| mINL-ELM | 30.93 (1,472) | .06 | **< .001** |  | 20.07 (1,472) | .04 | **< .001** |  | 7.25 (1,472) | .02 | **.007** |
| mELM-ISOS | 9.37 (1,472) | .02 | **.019** |  | 5.58 (1,472) | .01 | **.019** |  | 4.74 (1,472) | .01 | **.030** |
| mISOS-RPE | 19.39 (1,472) | .04 | **< .001** |  | 23.49 (1,472) | .05 | **< .001** |  | 29.81 (1,472) | .06 | **< .001** |
| ^a^ Adjusted for age, sex. | | | | | | | | | | | |

| **Supplementary Table 11.** Case-control differences in macular OCT measurements (photoreceptor layers), by subfield, Model 2 | | | | | | | | | | | |
| --- | --- | --- | --- | --- | --- | --- | --- | --- | --- | --- | --- |
| **Layer** | **Central subfield** ^a^ | | |  | **Inner subfield** ^a^ | | |  | **Outer subfield** ^a^ | | |
|  | *F* (df) | η²p | *p* |  | *F* (df) | η²p | *p* |  | *F* (df) | η²p | *p* |
| mINL-RPE | 30.03 (1, 407) | .07 | **< .001** |  | 25.65 (1, 407) | .06 | **< .001** |  | 18.75 (1, 407) | .04 | **< .001** |
| mINL-ELM | 22.50 (1, 407) | .05 | **< .001** |  | 16.54 (1, 407) | .04 | **< .001** |  | 6.64 (1, 407) | .02 | **.016** |
| mELM-ISOS | 4.61 (1, 407) | .01 | **.032** |  | 2.01 (1, 407) | .01 | .149 |  | 1.20 (1, 407) | .00 | .273 |
| mISOS-RPE | 10.18 (1, 407) | .02 | **.002** |  | 11.06 (1, 407) | .03 | **< .001** |  | 15.18 (1, 407) | .04 | **< .001** |
| ^a^ Adjusted for age, sex, Townsend deprivation index, body mass index, best corrected visual acuity, corneal hysteresis. | | | | | | | | | | | |

***OCT measurements in photoreceptor layers (by diagnosis)***

**Supplementary Tables 12–15** summarize case-control differences in macular OCT layers across PSD subgroups.

In the SZ subgroup, five layers showed significant group differences in Model 1: mRetina (*p* < .001), mINL-RPE (*p* < .001), mINL-ELM (*p* = .002), mISOS-RPE (*p* < .001), and mRPE (*p* = .009). In Model 2, mRetina (*p* = .002), mINL-RPE (*p* = .009), and mISOS-RPE (*p* = .021) remained significant. None of these layers remained significant after excluding individuals with cardiometabolic disorders (CMD), although mISOS-RPE approached significance (*p* = .050).

In the BD subgroup, six layers were significant in Model 1: mRetina (*p* < .001), mGC-IPL (*p* = .002), mINL-RPE (*p* < .001), mINL-ELM (*p* = .008), mELM-ISOS (*p* = .032), and mISOS-RPE (*p* = .004). In Model 2, mRetina (*p* < .001), mGC-IPL (*p* = .005), mINL-RPE (*p* = .001), and mINL-ELM (*p* = .009) remained significant. After excluding participants with CMD, only mINL-ELM remained significant (*p* = .023).

In the BD-P subgroup, three layers showed significant differences in Model 1: mRetina (*p* = .008), mINL-RPE (*p* < .001), and mISOS-RPE (*p* < .001). All remained significant in Model 2. In the CMD-excluded analysis, all three remained significant. Additionally, mGC-IPL became significant after CMD exclusion (*p* = .013), despite being nonsignificant in Models 1 and 2. However, results should be interpreted cautiously due to the small number of BD-P cases (n = 25), which limits statistical power.

In the MDD-P subgroup, only mELM-ISOS was significant in Model 1 (*p* = .032) but did not remain significant in Model 2 or in the CMD-excluded model. mRetina reached significance after CMD exclusion (*p* = .045), despite being nonsignificant in Models 1 and 2. Due to the very limited number of cases (n = 11), findings in the MDD-P subgroup should be interpreted cautiously, as statistical power was limited.

| **Supplementary Table 12.** Case-control differences in macular OCT measurements, SZ subgroup | | | | | | | | | | | |
| --- | --- | --- | --- | --- | --- | --- | --- | --- | --- | --- | --- |
| **Layer** | **Model 1** ^a^ | | |  | **Model 2** ^b^ | | |  | **Subgroup without CMD** ^c^ | | |
|  | *F* (df) | η²p | *p* |  | *F* (df) | η²p | *p* |  | *F* (df) | η²p | *p* |
| mRetina | 12.18 (1, 339) | .04 | **< .001** |  | 10.00 (1, 287) | .03 | .**002** |  | 2.72 (1, 229) | .02 | .100 |
| mRNFL | 0.00 (1, 339) | .00 | .978 |  | 0.01 (1, 287) | .00 | .905 |  | 0.34 (1, 229) | .00 | .562 |
| mGC-IPL | 1.90 (1, 339) | .01 | .169 |  | 1.69 (1, 287) | .01 | .195 |  | 1.65 (1, 229) | .01 | .200 |
| mINL | 0.60 (1, 339) | .00 | .439 |  | 0.81 (1, 287) | .00 | .368 |  | 0.38 (1, 229) | .00 | .541 |
| mINL-RPE | 26.84 (1, 339) | .07 | **< .001** |  | 6.87 (1, 287) | .03 | **.009** |  | 2.38 (1, 229) | .01 | .124 |
| mINL-ELM | 9.29 (1, 339) | .03 | **.002** |  | 2.88 (1, 287) | .01 | .091 |  | 0.54 (1, 229) | .00 | .464 |
| mELM-ISOS | 2.17 (1, 339) | .01 | .142 |  | 0.03 (1, 287) | .00 | .855 |  | 0.00 (1, 229) | .00 | .957 |
| mISOS-RPE | 23.71 (1, 339) | .07 | **< .001** |  | 5.46 (1, 287) | .02 | **.021** |  | 3.61 (1, 229) | .02 | **.050** |
| mRPE | 6.98 (1, 339) | .02 | **.009** |  | 0.17 (1, 287) | .01 | .173 |  | 3.39 (1, 229) | .02 | .067 |
| ^a^ Adjusted for age and sex.  ^b^ Adjusted for age, sex, Townsend deprivation index, body mass index, best corrected visual acuity, corneal hysteresis.  ^c^ This was 28 individuals with SZ and 209 healthy controls. Adjustment is the same as for model 2. | | | | | | | | | | | |

| **Supplementary Table 13.** Case-control differences in macular OCT measurements, BD subgroup | | | | | | | | | | | |
| --- | --- | --- | --- | --- | --- | --- | --- | --- | --- | --- | --- |
| **Layer** | **Model 1** ^a^ | | |  | **Model 2** ^b^ | | |  | **Subgroup without CMD** ^c^ | | |
|  | *F* (df) | η²p | *p* |  | *F* (df) | η²p | *p* |  | *F* (df) | η²p | *p* |
| mRetina | 18.07 (1, 331) | .05 | **< .001** |  | 11.54 (1, 286) | .03 | **< .001** |  | 2.26 (1, 219) | .01 | .134 |
| mRNFL | 1.36 (1, 331) | .00 | .245 |  | 0.54 (1, 286) | .00 | .465 |  | 0.32 (1, 219) | .00 | .575 |
| mGC-IPL | 9.43 (1, 331) | .03 | **.002** |  | 7.83 (1, 286) | .03 | **.005** |  | 1.98 (1, 219) | .01 | .161 |
| mINL | 1.88 (1, 331) | .01 | .172 |  | 0.90 (1, 286) | .00 | .344 |  | 0.21 (1, 219) | .00 | .651 |
| mINL-RPE | 16.60 (1, 331) | .05 | **< .001** |  | 10.74 (1, 286) | .04 | **.001** |  | 3.40 (1, 219) | .02 | .067 |
| mINL-ELM | 7.09 (1, 331) | .02 | **.008** |  | 7.40 (1, 286) | .03 | **.007** |  | 5.21 (1, 219) | .02 | **.023** |
| mELM-ISOS | 4.66 (1, 331) | .01 | **.032** |  | 1.22 (1, 286) | .00 | .270 |  | 0.20 (1, 219) | .00 | .658 |
| mISOS-RPE | 8.41 (1, 331) | .03 | **.004** |  | 2.62 (1, 286) | .01 | .107 |  | 0.20 (1, 219) | .00 | .887 |
| mRPE | 0.03 (1, 331) | .00 | .853 |  | 0.04 (1, 286) | .00 | .846 |  | 0.87 (1, 219) | .00 | .352 |
| ^a^ Adjusted for age and sex.  ^b^ Adjusted for age, sex, Townsend deprivation index, body mass index, best corrected visual acuity, corneal hysteresis.  ^c^ This was 18 individuals with BD and 209 healthy controls. Adjustment is the same as for model 2. | | | | | | | | | | | |

| **Supplementary Table 14.** Case-control differences in macular OCT measurements, BD-P subgroup | | | | | | | | | | | |
| --- | --- | --- | --- | --- | --- | --- | --- | --- | --- | --- | --- |
| **Layer** | **Model 1** ^a^ | | |  | **Model 2** ^b^ | | |  | **Subgroup without CMD** ^c^ | | |
|  | *F* (df) | η²p | *p* |  | *F* (df) | η²p | *p* |  | *F* (df) | η²p | *p* |
| mRetina | 7.25 (1, 259) | .03 | **.008** |  | 7.47 (1, 225) | .03 | **.007** |  | 10.55 (1, 210) | .05 | **.001** |
| mRNFL | 0.64 (1, 259) | .00 | .423 |  | 0.27 (1, 225) | .00 | .602 |  | 1.69 (1, 210) | .01 | .195 |
| mGC-IPL | 1.17 (1, 259) | .01 | .280 |  | 2.34 (1, 225) | .00 | .127 |  | 6.28 (1, 210) | .03 | **.013** |
| mINL | 0.39 (1, 259) | .00 | .553 |  | 0.68 (1, 225) | .00 | .409 |  | 2.93 (1, 210) | .01 | .088 |
| mINL-RPE | 12.37 (1, 259) | .05 | **< .001** |  | 10.74 (1, 225) | .04 | **.001** |  | 7.49 (1, 210) | .03 | **.007** |
| mINL-ELM | 1.70 (1, 259) | .01 | .194 |  | 7.40 (1, 225) | .03 | **.007** |  | 0.95 (1, 210) | .00 | .331 |
| mELM-ISOS | 0.19 (1, 259) | .00 | .666 |  | 1.22 (1, 225) | .00 | .270 |  | 0.03 (1, 210) | .00 | .861 |
| mISOS-RPE | 21.82 (1, 259) | .08 | **< .001** |  | 17.85 (1, 225) | .07 | **< .001** |  | 14.02 (1, 210) | .06 | **< .001** |
| mRPE | 1.38 (1, 259) | .01 | .241 |  | 0.60 (1, 225) | .00 | .438 |  | 4.56 (1, 210) | .02 | **.034** |
| ^a^ Adjusted for age and sex.  ^b^ Adjusted for age, sex, Townsend deprivation index, body mass index, best corrected visual acuity, corneal hysteresis.  ^c^ This was 9 individuals with BD-P and 209 healthy controls. Adjustment is the same as for model 2. | | | | | | | | | | | |

| **Supplementary Table 15.** Case-control differences in macular OCT measurements, MDD-P subgroup | | | | | | | | | | | |
| --- | --- | --- | --- | --- | --- | --- | --- | --- | --- | --- | --- |
| **Layer** | **Model 1** ^a^ | | |  | **Model 2** ^b^ | | |  | **Subgroup without CMD** ^c^ | | |
|  | *F* (df) | η²p | *p* |  | *F* (df) | η²p | *p* |  | *F* (df) | η²p | *p* |
| mRetina | 0.07 (1, 245) | .00 | .790 |  | 0.27 (1, 212) | .00 | .604 |  | 4.08 (1, 202) | .02 | **.045** |
| mRNFL | 0.20 (1, 245) | .00 | .653 |  | 1.07 (1, 212) | .01 | .302 |  | 1.05 (1, 202) | .01 | .308 |
| mGC-IPL | 0.92 (1, 245) | .00 | .339 |  | 0.70 (1, 212) | .00 | .403 |  | 2.24 (1, 202) | .01 | .136 |
| mINL | 0.16 (1, 245) | .00 | .905 |  | 0.10 (1, 212) | .00 | .755 |  | 2.14 (1, 202) | .01 | .145 |
| mINL-RPE | 0.40 (1, 245) | .00 | .529 |  | 0.14 (1, 212) | .00 | .707 |  | 2.24 (1, 202) | .01 | .136 |
| mINL-ELM | 0.01 (1, 245) | .00 | .931 |  | 0.02 (1, 212) | .00 | .890 |  | 1.74 (1, 202) | .01 | .189 |
| mELM-ISOS | 4.65 (1, 245) | .02 | **.032** |  | 3.43 (1, 212) | .02 | .065 |  | 0.22 (1, 202) | .00 | .640 |
| mISOS-RPE | 0.26 (1, 245) | .00 | .608 |  | 0.05 (1, 212) | .00 | .828 |  | 1.07 (1, 202) | .01 | .303 |
| mRPE | 0.72 (1, 245) | .00 | .396 |  | 1.21 (1, 212) | .01 | .273 |  | 1.13 (1, 202) | .01 | .289 |
| ^a^ Adjusted for age and sex.  ^b^ Adjusted for age, sex, Townsend deprivation index, body mass index, best corrected visual acuity, corneal hysteresis.  ^c^ This was 1 individual with MDD-P and 209 healthy controls. Adjustment is the same as for model 2. | | | | | | | | | | | |

***Association of retinal layer thickness with cognition markers***

***Supplementary Table 16*** summarizes cognitive variables significantly associated with retinal layer thickness. Fluid intelligence was positively associated with mRNFL thickness (p = .029), while prospective memory was associated with greater thickness in mINL-RPE (p = .017) and mELM-ISOS (p = .047). Additionally, fluid intelligence was significantly associated with mELM-ISOS thickness (p = .001), and processing speed was positively associated with mRPE thickness (p = .004). No other retinal layers showed significant associations with cognitive performance.

| **Supplementary** **Table 16.** Linear regression analyses identifying cognitive measurements associated with retinal layer thickness | | | | |
| --- | --- | --- | --- | --- |
|  | **β** | **B** | **95% CI** | ***p*** ^†^ |
| **mRetina** |  |  |  |  |
| Processing speed (*Reaction time test*) | -0.05 | -0.01 | (-0.02, 0.01) | .328 |
| Fluid intelligence (*Fluid intelligence test*) | 0.06 | 0.41 | (-0.19, 1.00) | .182 |
| Prospective memory (*Prospective memory test*) | 0.07 | 2.31 | (-0.93, 5.54) | .161 |
| **mRNFL** |  |  |  |  |
| Processing speed (*Reaction time test*) | -0.08 | -0.01 | (-0.01, 0.01) | .111 |
| Fluid intelligence (*Fluid intelligence test*) | 0.10 | 0.23 | (0.02, 0.43) | **.029** |
| Prospective memory (*Prospective memory test*) | 0.01 | 0.11 | (-0.88, 1.10) | .830 |
| **mGC-IPL** |  |  |  |  |
| Processing speed (*Reaction time test*) | 0.02 | 0.01 | (-0.01, 0.01) | .637 |
| Fluid intelligence (*Fluid intelligence test*) | -0.01 | -0.02 | (-0.28, 0.24) | .883 |
| Prospective memory (*Prospective memory test*) | 0.02 | 0.25 | (-1.09, 1.58) | .718 |
| **mINL** |  |  |  |  |
| Processing speed (*Reaction time test*) | -0.01 | 0.00 | (-0.01, 0.01) | .819 |
| Fluid intelligence (*Fluid intelligence test*) | -0.04 | -0.04 | (-0.14, 0.06) | .387 |
| Prospective memory (*Prospective memory test*) | -0.04 | -0.20 | (-0.73, 0.33) | .458 |
| **mINL-RPE** |  |  |  |  |
| Processing speed (*Reaction time test*) | -0.06 | -0.01 | (-0.01, 0.01) | .266 |
| Fluid intelligence (*Fluid intelligence test*) | 0.07 | 0.24 | (-0.08, 0.57) | .145 |
| Prospective memory (*Prospective memory test*) | 0.12 | 2.15 | (0.39, 3.92) | **.017** |
| **mINL-ELM** |  |  |  |  |
| Processing speed (*Reaction time test*) | -0.02 | -0.01 | (-0.01, 0.01) | .707 |
| Fluid intelligence (*Fluid intelligence test*) | 0.01 | 0.04 | (-0.23, 0.30) | .791 |
| Prospective memory (*Prospective memory test*) | 0.10 | 1.42 | (0.02, 2.83) | **.047** |
| **mELM-ISOS** |  |  |  |  |
| Processing speed (*Reaction time test*) | -0.03 | 0.01 | (-0.01, 0.01) | .534 |
| Fluid intelligence (*Fluid intelligence test*) | 0.16 | 0.11 | (0.04, 0.17) | **.001** |
| Prospective memory (*Prospective memory test*) | -0.01 | -0.03 | (-0.39, 0.33) | .864 |
| **mISOS-RPE** |  |  |  |  |
| Processing speed (*Reaction time test*) | -0.07 | -0.01 | (-0.01, 0.01) | .160 |
| Fluid intelligence (*Fluid intelligence test*) | 0.06 | 0.10 | (-0.10, 0.26) | .241 |
| Prospective memory (*Prospective memory test*) | 0.09 | 0.76 | (-0.12, 1.64) | .088 |
| **mRPE** |  |  |  |  |
| Processing speed (*Reaction time test*) | 0.14 | 0.01 | (0.01, 0.01) | .**004** |
| Fluid intelligence (*Fluid intelligence test*) | -0.09 | -0.12 | (-0.24, 0.01) | .055 |
| Prospective memory (*Prospective memory test*) | -0.07 | -0.44 | (-1.10, 0.21) | .185 |

| **Table S17.** Case-control differences in macular OCT measurements, repeated-measures ANCOVA, Age group with 40-55 years | | | | | | | | | | | |
| --- | --- | --- | --- | --- | --- | --- | --- | --- | --- | --- | --- |
| **Layer** | **Model 1** ^a^ | |  |  | **Model 2** ^b^ | |  |  | **Subgroup without CMD** ^c^ | | |
|  | *F* (df) | η²p | *p* |  | *F* (df) | η²p | *p* |  | *F* (df) | η²p | *p* |
| mRetina | 13.50 (1, 277) | .05 | **< .001** |  | 14.43 (1, 233) | .02 | **.041** |  | 3.89 (1, 156) | .03 | **.041** |
| mRNFL | 0.13 (1, 277) | .00 | .718 |  | 0.58 (1, 233) | .00 | .446 |  | 0.20 (1, 156) | .00 | .658 |
| mGC-IPL | 6.21 (1, 277) | .02 | **.013** |  | 1.57 (1, 233) | .01 | .211 |  | 3.86 (1, 156) | .02 | .051 |
| mINL | 3.58 (1, 277) | .01 | .060 |  | 0.61 (1, 233) | .00 | .435 |  | 0.81 (1, 156) | .01 | .369 |
| mINL-RPE | 16.78 (1, 277) | .06 | **< .001** |  | 4.23 (1, 233) | .02 | **.041** |  | 3.02 (1, 156) | .02 | .084 |
| mINL-ELM | 7.19 (1, 277) | .03 | **.008** |  | 2.10 (1, 233) | .01 | .149 |  | 1.83 (1, 156) | .01 | .178 |
| mELM-ISOS | 4.32 (1, 277) | .02 | **.038** |  | 2.37 (1, 233) | .01 | .125 |  | 0.10 (1, 156) | .00 | .756 |
| mISOS-RPE | 9.17 (1, 277) | .03 | **.003** |  | 1.21 (1, 233) | .01 | .273 |  | 1.13 (1, 156) | .01 | .290 |
| mRPE | 0.73 (1, 277) | .00 | .394 |  | 0.03 (1, 233) | .00 | .853 |  | 1.27 (1, 156) | .01 | .261 |
| ^a^ Adjusted for sex.  ^b^ Adjusted for sex, Townsend deprivation index, body mass index, best corrected visual acuity, spherical equivalent refractive error, corneal hysteresis.  ^c^ Consisted of 39 individuals with a psychosis-spectrum disorder and 125 healthy controls. Adjustment is the same as for model 2.  **Bold text**: Indicates significant measures. | | | | | | | | | | | |

| **Table S18.** Case-control differences in macular OCT measurements, repeated-measures ANCOVA, Age group with 56-70 years | | | | | | | | | | | |
| --- | --- | --- | --- | --- | --- | --- | --- | --- | --- | --- | --- |
| **Layer** | **Model 1** ^a^ | |  |  | **Model 2** ^b^ | |  |  | **Subgroup without CMD** ^c^ | | |
|  | *F* (df) | η²p | *p* |  | *F* (df) | η²p | *p* |  | *F* (df) | η²p | *p* |
| mRetina | 10.04 (1, 193) | .05 | **.002** |  | 9.92 (1, 161) | .06 | **.002** |  | 0.70 (1, 89) | .01 | .407 |
| mRNFL | 0.57 (1, 193) | .00 | .452 |  | 0.01 (1, 161) | .00 | .928 |  | 0.13 (1, 89) | .00 | .723 |
| mGC-IPL | 0.89 (1, 193) | .01 | .346 |  | 1.87 (1, 161) | .01 | .173 |  | 0.01 (1, 89) | .00 | .917 |
| mINL | 0.10 (1, 193) | .00 | .748 |  | 0.28 (1, 161) | .00 | .600 |  | 0.02 (1, 89) | .00 | .890 |
| mINL-RPE | 20.06 (1, 193) | .09 | **< .001** |  | 19.43 (1, 161) | .10 | **< .001** |  | 2.72 (1, 89) | .03 | .102 |
| mINL-ELM | 4.77 (1, 193) | .02 | **.030** |  | 6.18 (1, 161) | .04 | **.014** |  | 0.19 (1, 89) | .00 | .662 |
| mELM-ISOS | 1.53 (1, 193) | .01 | .218 |  | 2.07 (1, 161) | .00 | .451 |  | 0.10 (1, 89) | .00 | .748 |
| mISOS-RPE | 24.57 (1, 193) | .11 | **< .001** |  | 19.31 (1, 161) | .11 | **< .001** |  | 6.22 (1, 89) | .07 | **.014** |
| mRPE | 1.90 (1, 193) | .01 | .169 |  | 1.00 (1, 161) | .01 | .319 |  | 0.68 (1, 89) | .01 | .412 |
| ^a^ Adjusted for sex.  ^b^ Adjusted for sex, Townsend deprivation index, body mass index, best corrected visual acuity, spherical equivalent refractive error, corneal hysteresis.  ^c^ Consisted of 16 individuals with a psychosis-spectrum disorder and 81 healthy controls. Adjustment is the same as for model 2.  **Bold text**: Indicates significant measures. | | | | | | | | | | | |

***Prospective Memory and INL-RPE thickness***

Individuals with impaired prospective memory were significantly more likely to exhibit PSD+ (β=-0.16, B=-0.19, 95% CI [-0.29, -0.08], p<.001). However, impaired prospective memory was not significantly associated with INL-RPE thickness after adjusting for PSD status (β=0.09, B=1.68, 95% CI [-0.04, 3.40], p=.056). In contrast, PSD+ was significantly associated with INL-RPE thinning when adjusting for prospective memory (β=-0.25, B=-4.07, 95% CI [-5.58, -2.57], p<.001). Mediation analysis using the Sobel test indicated a significant indirect effect of PSD in the relationship between prospective memory and INL-RPE thickness (B=2.85, SE=0.27, p=0.004), with an indirect effect estimate of 0.76.

Point effect: 0.76 **

PSD

-0.186 (0.055) ***	-4.073 (0.767) ***

Prospective Memory +

INL-RPE thickness

1.676 (0.875)

SOBEL test: B = 2.85, SE = 0.27, p = 0.004

***Prospective Memory and INL-ELM thickness***

Individuals with impaired prospective memory were significantly more likely to exhibit PSD+ (β=-0.16, B=-0.19, 95% CI [-0.29, -0.08], p<.001). However, impaired prospective memory was not significantly associated with INL-ELM thickness after adjusting for PSD status (β=0.09, B=1.32, 95% CI [-0.09, 2.73], p=.065). PSD+ was significantly associated with INL-ELM thinning when adjusting for prospective memory (β=-0.15, B=-1.86, 95% CI [-3.09, -0.63], p=.003). Mediation analysis using the Sobel test indicated a significant indirect effect of PSD in the relationship between prospective memory and INL-ELM thickness (B=2.23, SE=0.15, p=0.025), with an indirect effect estimate of 0.35.

Point effect: 0.35 *

PSD

-0.186 (0.055) ***	-1.857 (0.626) ***

Prospective Memory +

INL-ELM thickness

1.320 (0.715)

SOBEL test: B = 2.23, SE = 0.15, p = 0.025

***Townsend Deprivation Index and INL-RPE thickness***

A greater Townsend Deprivation Index was significantly associated with an increased likelihood of PSD+ (β = 0.28, B = 0.04, 95% CI [0.03, 0.05], p < .001). Moreover, higher deprivation was significantly associated with INL-RPE thinning when adjusting for PSD status (β = -0.13, B = -0.30, 95% CI [-0.52, -0.09], p = .006). PSD+ was also significantly associated with INL-RPE thinning when adjusting for Townsend Deprivation Index (β = -0.23, B = -3.62, 95% CI [-5.07, -2.17], p < .001). Mediation analysis revealed a significant indirect effect of PSD in the relationship between Townsend Deprivation Index and INL-RPE thickness (Sobel test: B = -3.80, SE = 0.04, p < .001), with an indirect effect estimate of -1.52.

Point effect: -1.52 ***

PSD

0.04 (0.01) ***	-3.62 (0.74) ***

Townsend deprivation

INL-RPE thickness

-0.30 (0.11) **

SOBEL test: B = -3.80, SE = 0.04, p < 0.001
